# Exploratory electronic health record analysis with ehrapy

**DOI:** 10.1101/2023.12.11.23299816

**Authors:** Lukas Heumos, Philipp Ehmele, Tim Treis, Julius Upmeier zu Belzen, Altana Namsaraeva, Nastassya Horlava, Vladimir A. Shitov, Xinyue Zhang, Luke Zappia, Rainer Knoll, Niklas J. Lang, Leon Hetzel, Isaac Virshup, Lisa Sikkema, Eljas Roellin, Fabiola Curion, Roland Eils, Herbert B. Schiller, Anne Hilgendorff, Fabian J. Theis

## Abstract

With progressive digitalization of healthcare systems worldwide, large-scale collection of electronic health records (EHRs) has become commonplace. However, an extensible framework for comprehensive exploratory analysis that accounts for data heterogeneity is missing. Here, we introduce ehrapy, a modular open-source Python framework designed for exploratory end-to-end analysis of heterogeneous epidemiology and electronic health record data. Ehrapy incorporates a series of analytical steps, from data extraction and quality control to the generation of low-dimensional representations. Complemented by rich statistical modules, ehrapy facilitates associating patients with disease states, differential comparison between patient clusters, survival analysis, trajectory inference, causal inference, and more. Leveraging ontologies, ehrapy further enables data sharing and training EHR deep learning models paving the way for foundational models in biomedical research. We demonstrated ehrapy’s features in five distinct examples: We first applied ehrapy to stratify patients affected by unspecified pneumonia into finer-grained phenotypes. Furthermore, we revealed biomarkers for significant differences in survival among these groups. Additionally, we quantify medication-class effects of pneumonia medications on length of stay. We further leveraged ehrapy to analyze cardiovascular risks across different data modalities. Finally, we reconstructed disease state trajectories in SARS-CoV-2 patients based on imaging data. Ehrapy thus provides a framework that we envision will standardize analysis pipelines on EHR data and serve as a cornerstone for the community.

## Introduction

Collections of clinical and epidemiological data, demographic profiles, pharmacological records, biobank data, and more, called electronic health records (EHR), are becoming commonplace owing to increasingly standardized data collection^1^ and digitalization in healthcare institutions. EHRs collected at medical care sites serve as efficient storage and sharing units of health information^2^ enabling the informed treatment of individuals using the patient’s complete history^3^. Routinely collected EHR data is approaching genomic scale in size and complexity^4^, leading to challenges in extracting information from these data without quantitative analysis approaches. Application of such approaches to EHR databases^1,5–9^ has enabled the prediction and classification of disease^10,11^, study population health^12^, determine optimal treatment policies^13,14^, simulate clinical trials^15^, and stratify patients^16^.

However, current EHR datasets suffer from severe limitations that impact analysis quality, ranging from data collection issues and data inconsistency to data diversity. EHR data collection and sharing problems often arise due to non-standardized formats, with disparate systems using exchange protocols such as Health Level Seven International (HL7) and Fast Healthcare Interoperability Resources (FHIR)^17^. In addition, EHR data is stored in various on-disk formats including, but not limited to, relational databases, CSV, XML, and JSON formats. These variations pose challenges with respect to data retrieval, scalability, interoperability, and data sharing. EHR data can further be representationally and semantically inconsistent when data varies in format, unit, granularity and measurement protocol. In such cases, the usage of multiple or non-standard medical coding schemes between datasets prevents data sharing and integration. Data may also contradict itself, such as when measurements were recorded for deceased patients^18,19^. Finally, technical variation and different data collection standards result in distribution differences between datasets^20,21^. The diversity of EHR data that comprises demographics, laboratory results, vital signs, diagnoses, medications, X-rays, written notes, and even omics measurements amplifies all aforementioned issues, further increasing the challenge of comparing and integrating datasets.

To adequately address these obstacles, the usage of EHR data requires careful data preprocessing, presumed to account for 80% of the effort when performing a typical analysis or model development^22^. A few EHR data preprocessing and analysis workflows have been previously developed^4,23–27^, but none of them enable the analysis of heterogeneous data, provide in-depth documentation, are software packages or allow for exploratory visual analysis. Current EHR analysis pipelines therefore differ strongly in their approaches and are often commercial, vendor specific solutions^28^. This is in contrast to strategies using community standards for the analysis of omics data such as Bioconductor^29^ or scverse^30^. As a result, EHR data frequently remain underexplored and are commonly only investigated for a particular research question^31^. Even in such cases, EHR data are then frequently fed into machine learning models with severe data quality issues that greatly impact prediction performance and generalizability^32^.

To address this lack of analysis tooling, we developed the EHR Analysis in Python framework, ehrapy, a Python based EHR analysis framework that enables exploratory analysis of diverse EHR datasets. The ehrapy package is purpose-built to organize, analyze, visualize and statistically evaluate complex EHR data. Ehrapy’s versatility allows its application to datasets of different data types, sizes, diseases, and origins. To demonstrate this versatility, we apply ehrapy to datasets obtained from EHR and population based studies, including imaging: In the Paediatric Intensive Care EHR database^33^, we stratify patients diagnosed with “unspecified pneumonia” into distinct clinically relevant groups, extract clinical indicators of pneumonia through statistical analysis, and quantify medication-class effects on length of stay with causal inference.

Using the UK-Biobank^34^ (UKB), a population cohort comprising over 500,000 participants from the United Kingdom, we employ ehrapy to explore cardiovascular risk factors at population scale using clinical predictors, metabolomics, genomics, and retinal imaging-derived features. Finally, we focus on image analysis to project disease progression through fate mapping in patients affected with COVID-19 using chest X-rays. We provide online links to additional use cases that demonstrate ehrapy’s usage with further datasets, including MIMIC-II^35^, and for various medical conditions, such as patients subject to indwelling arterial catheter usage. Ehrapy is compatible with any EHR dataset that can be transformed into vectors and is accessible as a scalable, user-friendly open-source software package hosted at https://github.com/theislab/ehrapy and installable from PyPI and Conda. It comes with comprehensive documentation, tutorials, and further examples, available at https://ehrapy.org.

## Results

### Ehrapy: a framework for exploratory and targeted EHR data analysis

The foundation of ehrapy is a robust and scalable implementation-independent data storage backend which is combined with a series of preprocessing and analysis modules. In ehrapy, EHR data is organized as a data matrix where observations are individual patient visits (or patients, in the absence of follow-up visits), and variables represent all measured quantities (**Methods**). These data matrices are stored together with metadata of observations, variables, and unstructured annotations. By leveraging the open AnnData (annotated data) data structure that implements this design, ehrapy builds upon established, scalable standards and is compatible with analysis and visualization functions provided by the single-cell omics scverse^30^ ecosystem. Ehrapy thus allows for larger-than-memory analyses by leveraging HDF5-^36^ or Zarr-based^37^ AnnData objects. Both formats are platform, framework and programming language independent formats and readers are also available in R, Julia and Javascript^38^. We additionally provide a dataset module with more than 20 public loadable EHR datasets in AnnData format to kickstart analysis and development with ehrapy.

For standardized analysis of EHR data, it is crucial that these data are encoded and stored in consistent, reusable formats. Thus, ehrapy requires that input data is organized in structured vectors. Readers for common formats such as CSV, OMOP^39^, or SQL databases are available in ehrapy. Data loaded into AnnData objects can be mapped against hierarchical ontologies such as MONDO^40^, International Statistical Classification of Diseases (ICD)^41^, Human Phenotype Ontology^42^, or Phecodes^43^ (**Methods**). Clinical keywords of free text notes can be automatically extracted (**Methods**).

Powered by scanpy, which scales to hundreds of millions of observations^44^ and the machine learning library scikit-learn^45^, ehrapy provides more than 100 composable analysis functions organized in modules from which custom analysis pipelines can be built. In such pipelines, each function directly interacts with the AnnData object and adds all intermediate results to the object for simple access and reuse of information. To facilitate setting up these pipelines, ehrapy guides analysts through a general analysis pipeline (**Figure 1**) consisting of data preparation, data preprocessing and knowledge inference. At any step of an analysis pipeline, community software packages can be injected without any vendor lock-in. Since ehrapy is built on open standards, it can be purposefully extended to solve new challenges. Ehrapy natively interfaces with the scientific Python ecosystem via Pandas^46^ and Numpy^47^. The development of deep-learning models for EHR data^48^ is further accelerated through compatibility with pathml^49^, a unified framework for whole-slide image analysis in pathology, and scvi-tools^50^ which provides data loaders for loading tensors from AnnData objects into PyTorch^51^ or Jax arrays^52^ to facilitate the development of generalizing foundational models for medical artificial intelligence^53^.

**Figure 1.**
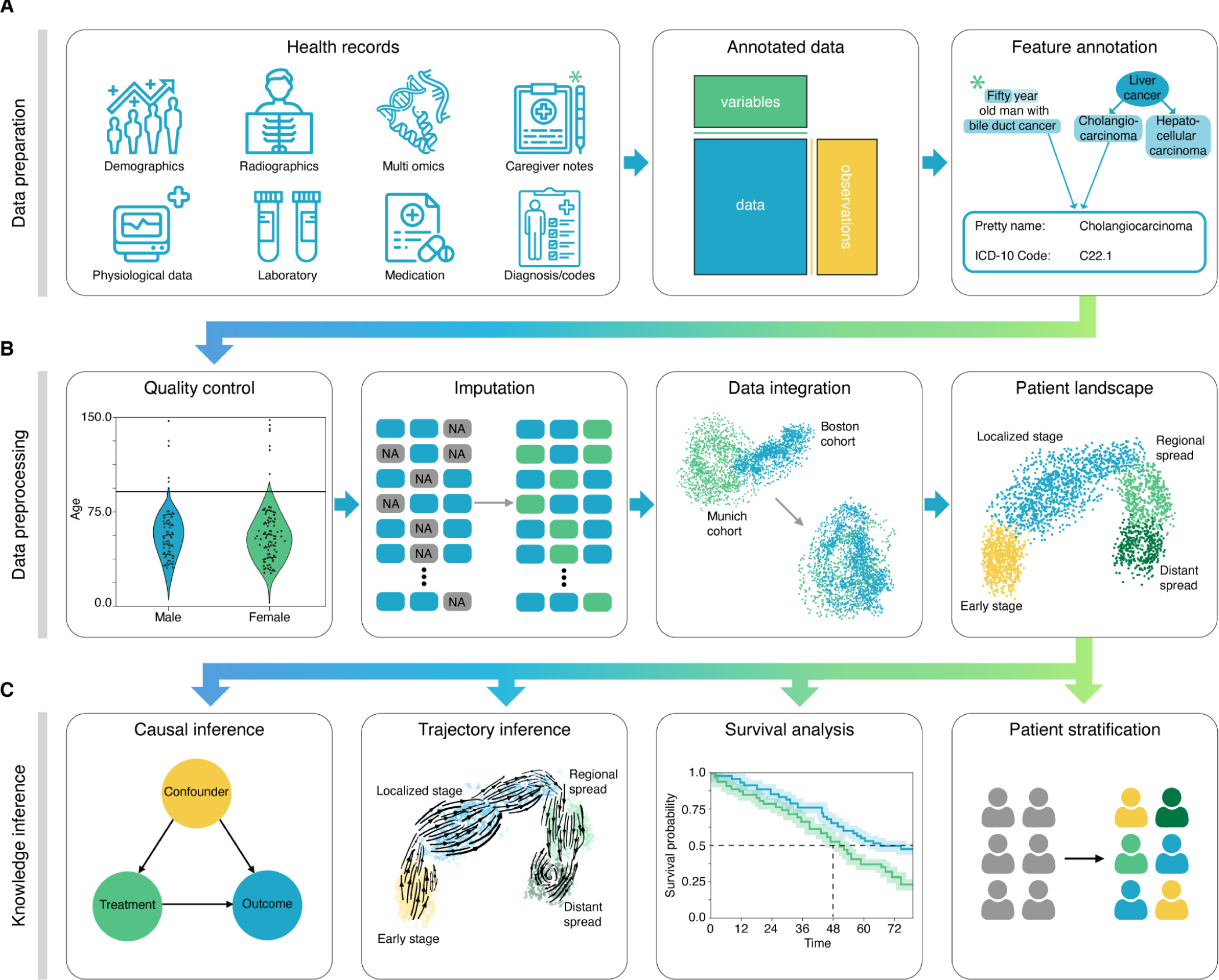
Schematic overview of ehrapy analysis. (A) Heterogeneous health data are first stored in memory as an Annotated Data (AnnData) object with patients or patient visits as observations and variables as columns. Next, the data are first mapped against ontologies and key terms are extracted from free text notes. (B) The EHR data are subject to quality control where impossible or spurious measurements are removed. Subsequently, categorical data are encoded and missing data are imputed. Data from different sources with data distribution shifts are integrated, embedded, clustered and annotated in a patient landscape. (C) Further downstream analyses depend on the question of interest and can include the inference of causal effects and trajectories, survival analysis, or patient stratification.

In the general ehrapy analysis pipeline, EHR data is first examined to detect quality issues by inspecting feature distributions (**Methods**). Strong outliers determined this way may represent erroneous measurements that could skew any calculations and can therefore be winsorized, clipped, or removed. Automatic quality control further detects visits and features with high missing rates that ehrapy can then impute (**Methods**). Subsequently, ehrapy’s normalization and encoding functions (**Methods**) are used to ensure a consistent numerical data representation that enables data integration while regressing out dataset shift effects (**Methods**). The size and complexity of EHR data requires lower dimensional representations that can subsequently be visualized, clustered and annotated to obtain a patient landscape (**Methods**). Such annotated groups of patients can be used for statistical comparisons to find differences in features among them to ultimately learn markers of patient states.

As analysis goals can differ between users and datasets, the general ehrapy analysis pipeline is opened up in the final knowledge inference step such that the right modules can be chosen to best answer the question at hand. Ehrapy provides advanced statistical methods for group comparison and linear models together with extensive support for survival analysis (**Methods**) enabling the discovery of biomarkers. Furthermore, ehrapy provides functions for causal inference to go from statistically determined associations to causal relations (**Methods**). Disease progression analysis represents a further analysis goal for EHR data that ehrapy also provides support for in the form of trajectory inference methods (**Methods**). Patient visits in aggregated EHR data can be regarded as snapshots, offering a limited perspective that may not encompass the entirety of a patient’s clinical picture or the progression of their disease. Individual measurements taken at specific time points might not adequately reflect the underlying progression of disease but result from unrelated variation due to, for example, day-to-day differences^54–56^. Therefore, disease progression models should rely on analysis of the underlying clinical data rather than of a notion of time as disease progression in an individual patient may not be monotonous in time. Ehrapy’s integration into the larger scverse ecosystem allows for the usage of advanced trajectory inference methods to facilitate novel views on EHR data through compatible tools like Diffusion Pseudotime^57^ or CellRank^58^. We show that these tools can order snapshots to calculate a pseudotime that can adequately reflect the progression of the underlying clinical process without requiring completeness of real-time measurements in all patients. Given a sufficient number of snapshots, ehrapy increases the potential to capture disease progression that is likely not robustly captured within a single EHR record, but rather across several.

### Ehrapy enables patient stratification in unspecified pneumonia cases

To demonstrate ehrapy’s capability to analyze heterogeneous datasets from a broad patient set across multiple care units, we applied our novel strategy to the Paediatric Intensive Care (PIC)^33^ database. The PIC database is a single-center, bilingual (English and Chinese) database hosting information on children admitted to critical care units at the Children’s Hospital of Zhejiang University School of Medicine in China. It contains 13,499 distinct hospital admissions of 12,881 individual pediatric patients admitted between 2010 and 2018 for which demographics, diagnoses, doctors’ notes, vital signs, laboratory and microbiology tests, medications, fluid balances, and more were collected (**Figure 2A, Supplementary Figure 1, Methods**). Following missing data imputation and subsequent preprocessing (**Figure 2B-C**, **Supplementary Figure 2, Methods**), we generated a Uniform Manifold Approximation and Projection (UMAP) embedding to visualize variation across all patients using ehrapy (**Figure 3A**). This visualization of a low dimensional patient manifold shows the heterogeneity of the collected data in the PIC database with malformations, perinatal, and respiratory being the most abundant ICD chapters (**Figure 3B**).

**Figure 2.**
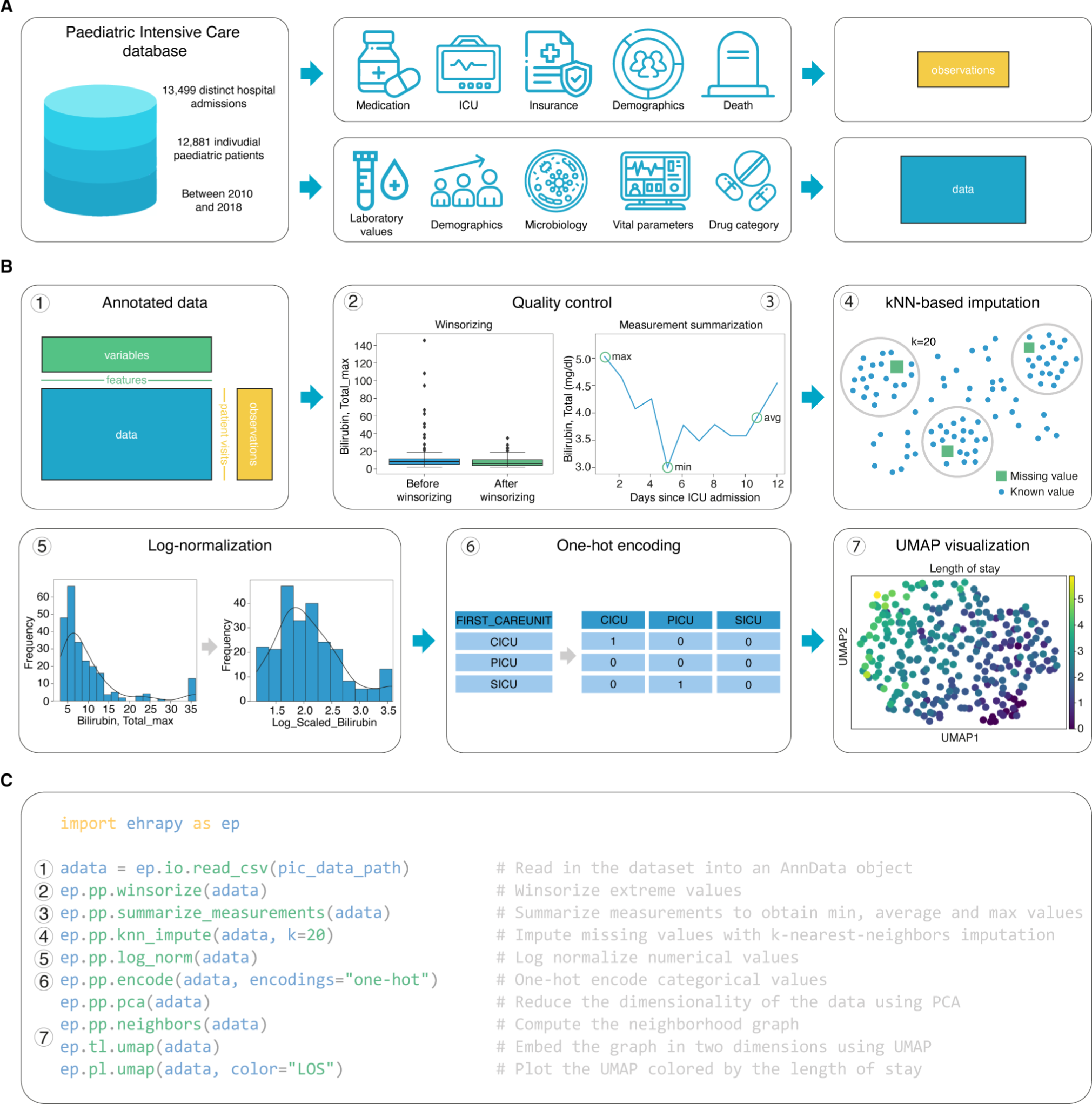
Preprocessing of the Paediatric Intensive Care (PIC) dataset with ehrapy. (A) Heterogeneous data of the PIC database was stored in “data” (matrix that is used for computations) and “observations” (metadata per patient). During quality control further annotations are added to the “variables” (metadata per feature) slot. (B) Preprocessing steps of the PIC dataset. (C) Minimal example of the function calls in the data analysis pipeline that resembles the preprocessing steps in (B) using ehrapy.

**Figure 3.**
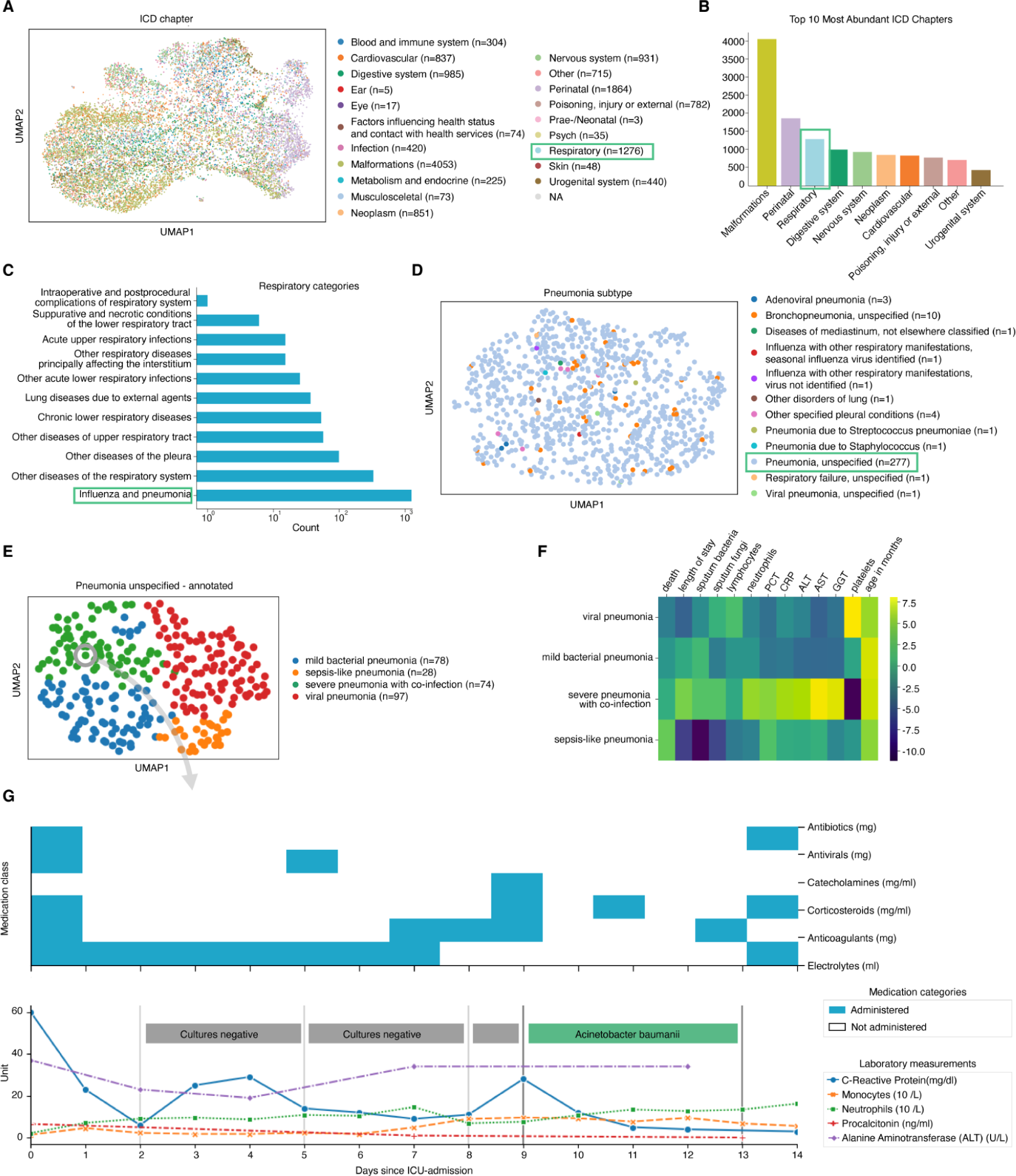
PIC dataset overview and annotation of patients diagnosed with unspecified pneumonia. (A) Uniform manifold approximation and projection (UMAP) of all patient visits in the intensive care unit (ICU) with primary discharge diagnosis grouped by ICD chapter. The organ heterogeneity in the malformation chapter prevents very distinct clusters per chapter. (B) The abundance and complexity of respiratory diseases encouraged us to investigate them further. (C) Respiratory categories show the abundance of influenza and pneumonia diagnoses that we investigated further. (D) We observed the “unspecified pneumonia” sub-group which led us to investigate and annotate it further in more detail. (D) The previously “unspecified pneumonia” labeled patients were annotated using several clinical features (**Supplementary Figure 3**) of which the most important ones are shown in the heatmap (E). (G) Example disease progression of an individual child with pneumonia illustrating pharmacotherapy over time until positive acinetobacter baumannii swab.

The most common respiratory disease categories (**Figure 3C**) were pneumonia and influenza (n=1229). We focused on pneumonia to apply ehrapy to a challenging, broad-spectrum disease across age. Pneumonia is a prevalent respiratory infection that poses a substantial burden on public health^59^ and is characterized by inflammation of the alveoli and distal airways^59^. Individuals with pre-existing chronic conditions are particularly vulnerable, as are children under the age of five^60^. Pneumonia can be caused by a range of microorganisms, encompassing bacteria, respiratory viruses, and fungi.

From these cases, we selected the age group “youths” (13 months to 18 years of age) for further analysis, thereby addressing a total of 265 patients. Neonates (0 to 28 days) and infants (28 days to 13 months old) were excluded from the analysis as the disease context is significantly different in these age groups due to distinct anatomical and physical conditions. These 265 patients dominated the group of pneumonia cases and were diagnosed with “unspecified pneumonia” by the collectors of the original database (**Figure 3D**).

Patients were 61% male, had a total of 277 admissions, a mean age at admission of 54 months (median of 38 months) and an average length of stay (LOS) of 15 days, with a median of 7 days. Of these, 152 patients were admitted to the pediatric ICU (PICU), 118 to the general ICU (GICU), four to the surgical ICU (SICU), and three to the cardiac ICU (CICU). Laboratory measurements generally contained between 12% and 14% missing data with serum procalcitonin (PCT) (24.5%) and C-reactive protein (CRP) (16.8%) being notable exceptions. Parameters assigned as “vital signs” contained between 44% and 54% missing values. Stratifying patients with unspecified pneumonia further enables a more nuanced understanding of the disease, potentially facilitating tailored approaches to treatment.

To deepen clinical phenotyping for the disease group “unspecified pneumonia”, we calculated a k-nearest-neighbor graph to cluster patients into subgroups and visualize these in UMAP space, thereby progressing to the next step in ehrapy’s workflow (**Figure 1, Methods**). Leiden clustering^61^ identified four patient groupings with distinct clinical features that we annotated (**Figure 3E**).

To identify the laboratory values, medications, and pathogens that were most characteristic for these four groups (**Figure 3F**), we applied t-tests for numerical and g-tests for categorical data using ehrapy (**Supplementary Figure 3, Methods**). Based on this analysis, we identified patient subgroups with “sepsis-like, “severe pneumonia with co-infection”, “viral pneumonia” and “mild pneumonia” phenotypes.

The “sepsis-like” group of patients (n=28) were characterized by rapid disease progression as exemplified by an increased number of deaths (adj. P≤ 5.04e-03), indication of multiple organ failure such as elevated creatinine (adj. P ≤ 0.01) or reduced albumin levels (adj. P ≤ 2.89e-04) together with increased expression levels and peaks of inflammation markers including PCT (adj. P ≤ 3.01e-02), whole blood cell count, neutrophils, lymphocytes, monocytes, lower platelet counts (adj. P ≤ 6.3e-02) and changes in electrolyte levels, i.e. lower potassium levels (adj. P ≤ 0.09e-02). Patients we associated with the term “severe pneumonia with co-infection” (n=74) were characterized by prolonged ICU stays (adj. P≤ 3.59e-04), organ affection such as higher levels of creatinine (adj. P ≤ 1.10e-04), increased inflammation markers such as peaks of PCT (adj. P≤ 5.06e-05), CRP (adj. P≤ 1.40e-06), and neutrophils (adj. P≤ 8.51e-06), together with valleys of lower platelet count (adj. P ≤ 5.40e-23), detection of bacteria in combination with additional pathogens such as fungals in sputum samples (adj. P≤ 1.67e-02), and increased application of medication including antifungals (adj. P≤ 1.30e-04) and catecholamines (adj. P≤ 2.0e-02)). Patients in the “mild pneumonia” cluster were characterized by positive sputum cultures in the presence of relatively lower inflammation markers such as PCT (adj. P≤ 1.63e-03) and CRP (adj. P≤ 0.03e-01), while receiving antibiotics more frequently (adj. P ≤ 1.00e-05), and additional medications (electrolytes, blood thinners and circulation supporting medications) (adj P≤ 1.00e-05). Finally, patients in the“viral infection” cluster were characterized by shorter lengths of stays (adj. P ≤ 8.00e-06), a lack of non-viral pathogen detection in combination with higher lymphocyte counts (adj. P≤ 0.01 avg) and peaks of PCT (adj. P ≤ 0.03e-02), reduced application of catecholamines (adj. P≤ 5.96e-07), antibiotics (adj. P≤ 8.53e-06), antifungals (adj. P≤ 5.96e-07).

To demonstrate the ability of ehrapy to examine EHR data from different levels of resolution, we additionally reconstructed a case from the “severe pneumonia with co-infection” group (**Figure 3G**). In this case, the analysis revealed that CRP levels remained elevated despite broad spectrum antibiotic treatment until a positive “Acinetobacter baumanii” result led to a change in medication and a subsequent decrease in CRP and monocyte levels.

### Ehrapy empowers extraction of clinical indicators for unspecified pneumonia

Ehrapy’s survival analysis module allowed us to identify clinical indicators of disease or disease stages that could be used as biomarkers through Kaplan-Meier analysis. We found strong variance in overall aspartate aminotransferase (AST), alanine aminotransferase (ALT), and gamma-glutamyl transferase (GGT) levels (**Figure 4A**) including changes over time (**Supplementary Figure 4A-B**) in all four “unspecified pneumonia” groups. Routinely used to assess liver function, studies provide evidence that AST, ALT, GGT levels are elevated during respiratory infections^62,63^ including severe pneumonia ^64^, and can guide diagnosis and management of pneumonia in children^62^.

**Figure 4.**
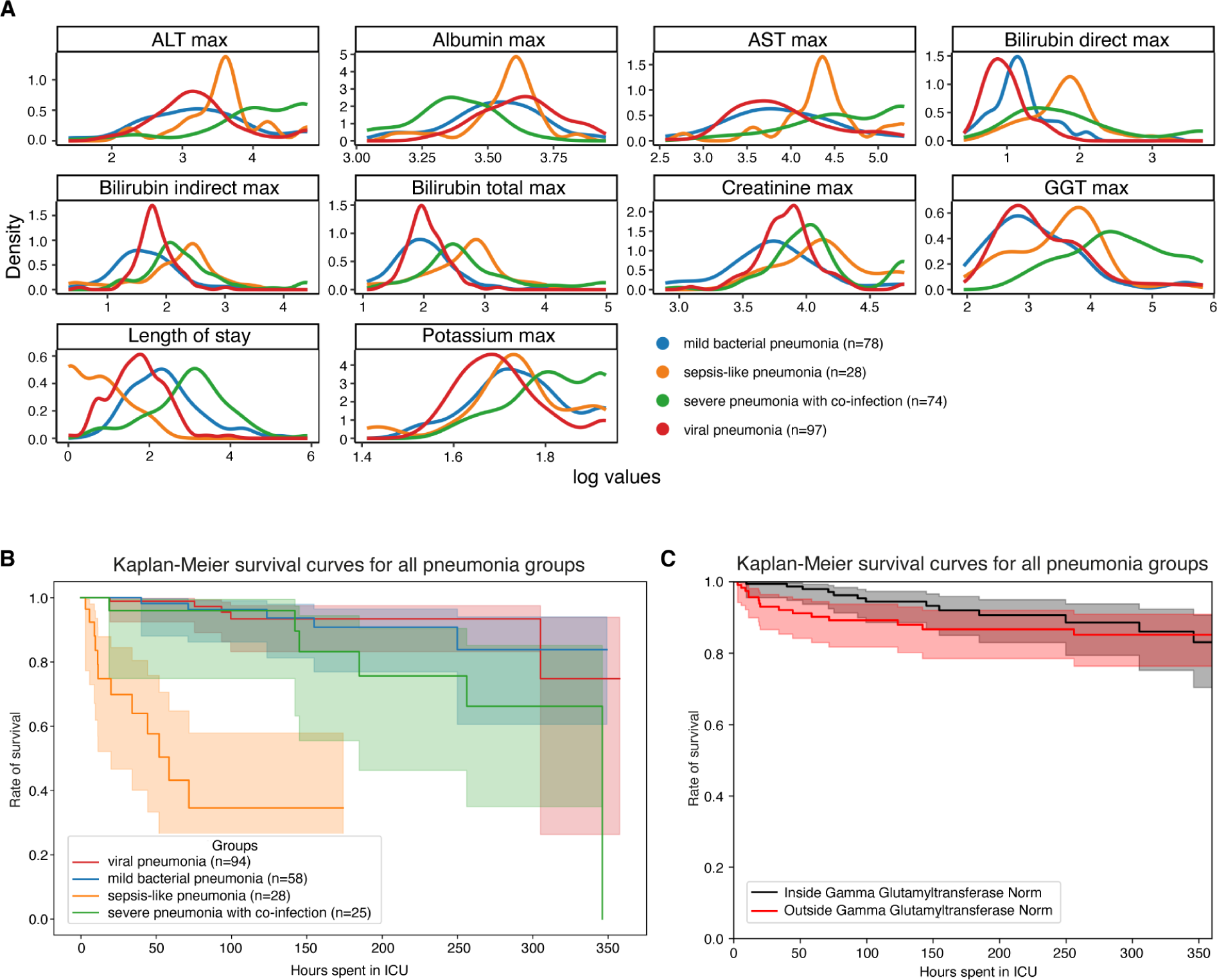
Survival analysis of patients diagnosed with unspecified pneumonia. (A) Line plots of major hepatic system lab measurements per group show variance in the measurements per pneumonia group. (B) Kaplan-Meier survival curves demonstrate lower survival for “sepsis-like” and “severe pneumonia with co-infection” groups. (C) Kaplan-Meier survival curves for children with Gamma Glutamyltransferase measurements outside the norm range display lower survival.

We confirmed reduced survival in more severely affected children (“sepsis-like pneumonia” and “severe pneumonia with co-infection”) using Kaplan-Meier curves and a multivariate log-rank test (**Figure 4B**; P ≤1.09e-18) through ehrapy. To verify the association of this trajectory with altered AST, ALT, GGT expression levels, we further grouped all patients based on liver enzyme reference ranges (**Methods, Table 1**). By Kaplan-Meier survival analysis, cases with peaks of GGT (P ≤ 1.4e-02), ALT (P ≤ 2.9e-02), and AST (P ≤ 4.8e-04) in “outside the norm” were found to correlate with lower survival in all groups (**Figure 4C, Supplementary Figure 4**), in line with previous studies^62,63^.

**Table 1.** Reference ranges of liver markers.

| Measurement | Minimum | Maximum |
| --- | --- | --- |
| Aspartate aminotransferase (AST) | 10 IU/L | 40 IU/L |
| Alanine transaminase (ALT) | 10 IU/L | 35 IU/L |
| Gamma-glutamyl transferase (GGT) | 0 IU/L | 35 IU/L |

### Ehrapy allows for medication-class effect quantification of length of stay by causal inference

Pneumonia requires case-specific medications due to its diverse causes. To demonstrate the potential of ehrapy’s causal inference module, we quantified the effect of medication on ICU length of stay to evaluate case-specific administration of medication. In contrast to causal discovery that attempts to find a causal graph reflecting the causal relationships, causal inference is a statistical process used to investigate possible effects when altering a provided system, as represented by a causal graph and observational data (**Figure 5A**)^65^. This approach allows researchers to identify and quantify the impact of specific interventions or treatments on outcome measures, thereby providing insight for evidence-based decision-making in healthcare. Causal inference relies on datasets incorporating multiple perturbation conditions, commonly termed “interventions”, to accurately quantify effects.

**Figure 5.**
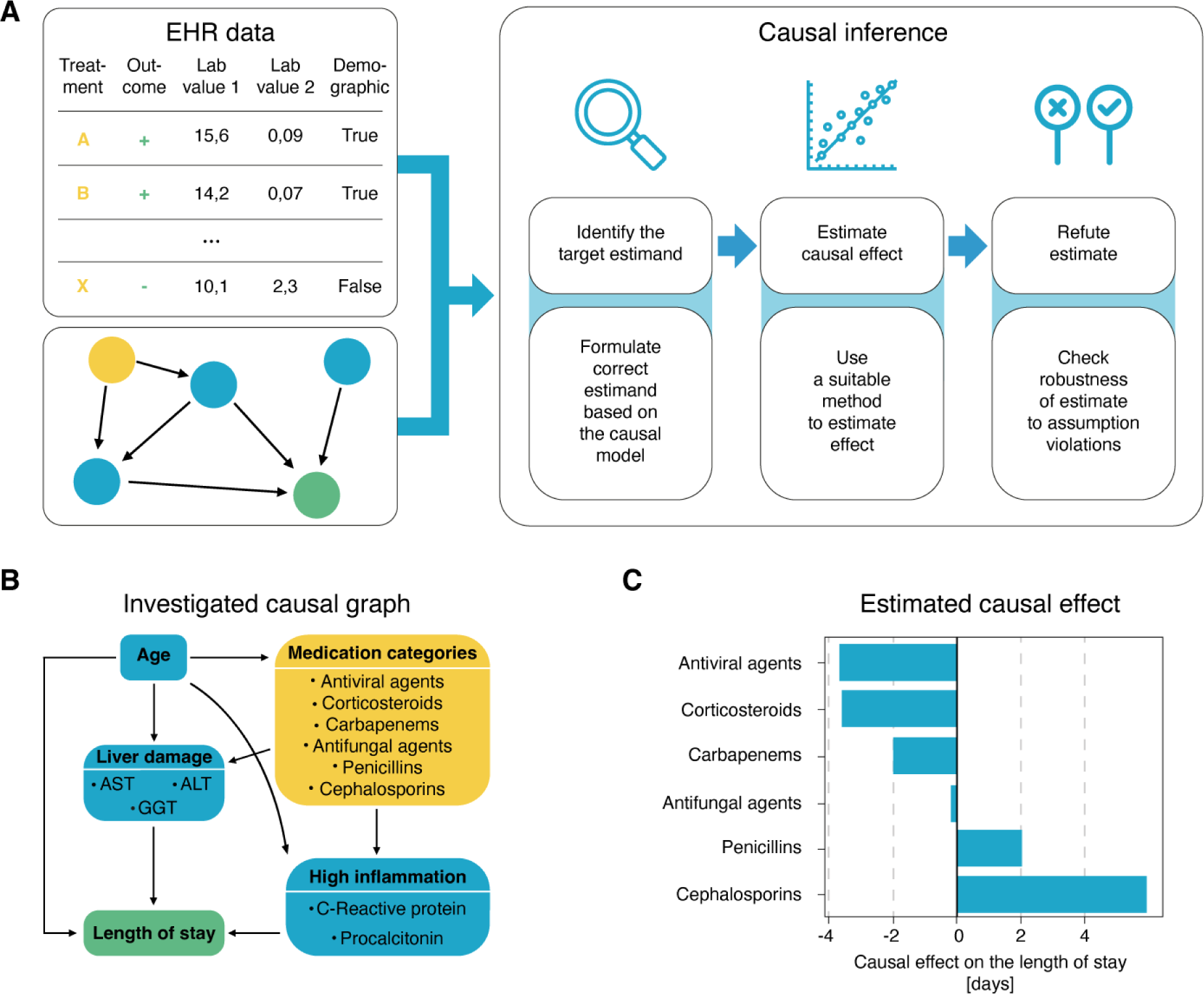
Causal inference of length of stay affected by different medication types. (A) ehrapy’s causal module is based on the strategy of the tool dowhy. Here, EHR data containing treatment, outcome, and measurements, and a causal graph serve as input for causal effect quantification. The process includes the identification of the target estimand based on the causal graph, the estimation of causal effects using various models, and finally refutation where sensitivity analyses and refutation tests are performed to assess the robustness of the results and assumptions. (B) Curated causal graph using age, liver damage, and inflammation markers as disease progression proxies together with medications as interventions to assess the causal effect on length of ICU stay. (C) Determined causal effect strength on length of stay in days of administered medication categories showcases lower expected length of stays in cases where corticosteroids, carbapenems, and antivirals are administered.

We manually constructed a minimal causal graph with ehrapy (**Figure 5B**) on records of treatment with corticosteroids, carbapenems, penicillins, cephalosporins, antifungal, and antiviral medication (**Supplementary figure 5)**. We used these as “interventions’’, and assumed the medications affect disease progression proxies such as inflammation markers and markers of organ function. The selection of “interventions” is consistent with current treatment standards for bacterial pneumonia and respiratory distress^66,67^.

Based on the approach of the tool dowhy^68^ (**Figure 5A**), ehrapy’s causal module identified the application of corticosteroids, antivirals, and carbapenems to be associated with shorter lengths of stay, in line with current evidence^60,69,70,71^. In contrast, penicillins and cephalosporins were associated with longer lengths of stay (**Figure 5C**), whereas antifungal medication did not strongly influence length of stay (**Figure 5c**).

### Ehrapy facilitates automated extraction of population-scale risk factors in the UK-Biobank

To demonstrate ehrapy’s scalability and to illustrate the advantages of using a unified data management and quality control framework like ehrapy when performing multimodal survival modeling, we modeled myocardial Infarction risk using Cox Proportional-Hazards Models on UK-Biobank (UKB)^34^ data. Large population cohort studies like the UK-Biobank enable the investigation of common diseases across a wide range of modalities including genomics, metabolomics, proteomics, imaging data, and common clinical variables (**Figure 6A-B**). From these, we used a publicly available polygenic risk score for coronary heart disease^72^ comprising 6.6 million variants, 80 nuclear magnetic resonance (NMR) spectroscopy-based metabolomics^73^ features, 81 features derived from retinal optical coherence tomography^74,75^, and the Framingham Risk Score^76^ feature set, which includes known clinical predictors such as age, sex, BMI, blood pressure, smoking behavior, and cholesterol levels. We filtered the feature space by excluding features with more than 10% missingness in the respective population and imputed the remaining missing values (**Methods**). Further, individuals with comorbidities were excluded from the analyses. We selected 29,216 individuals for whom all of these data types were available (**Supplementary Figure 6**-**7, Methods**). Myocardial Infarction, as defined by our mapping to the Phecode nomenclature^43^, was defined as the endpoint (**Figure 6C**). We modeled the risk for myocardial Infarction one year after either the metabolomic sample was obtained or imaging was performed.

**Figure 6.**
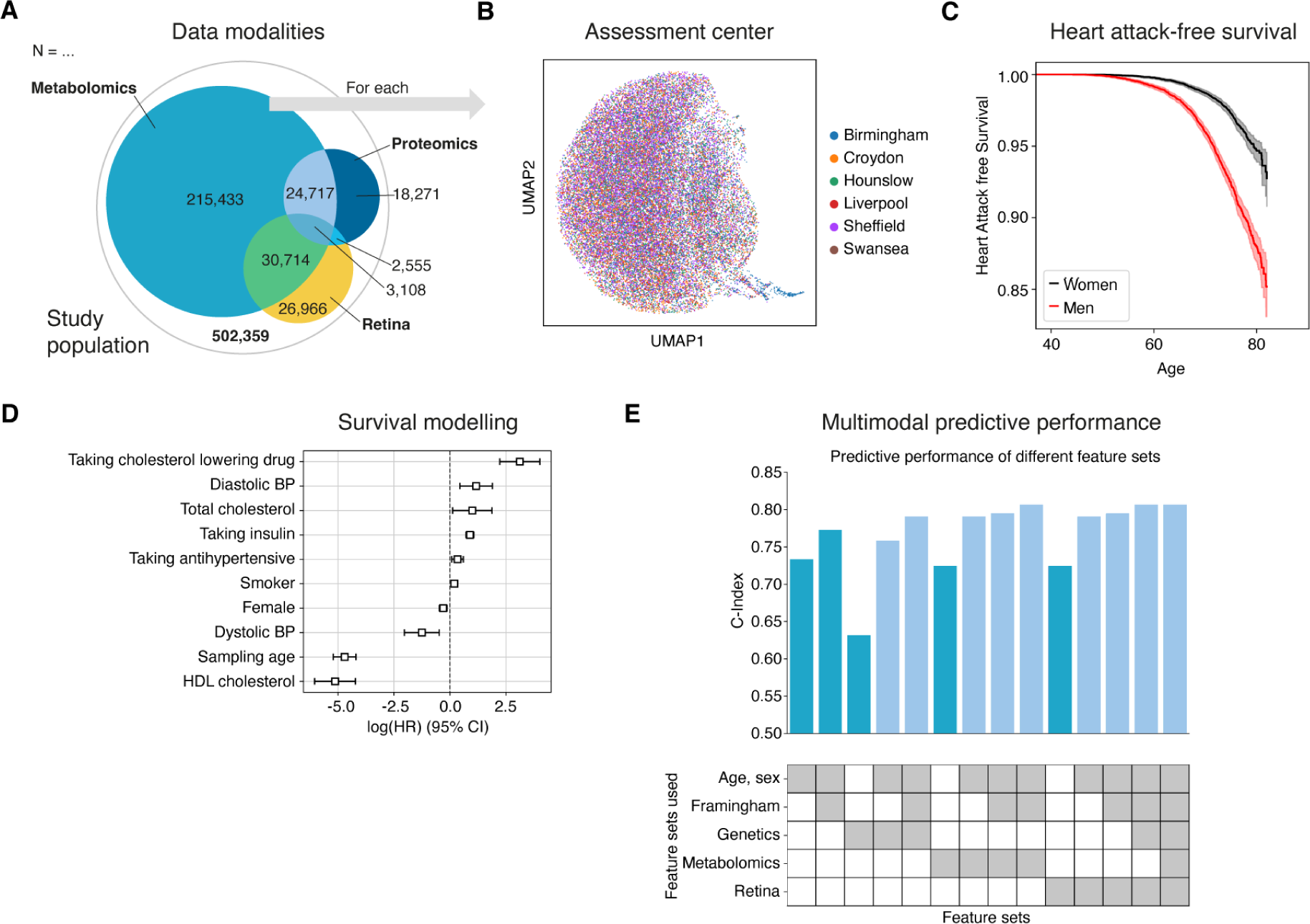
Analysis of myocardial Infarction risk in the UK-Biobank. (A) The UK-Biobank includes 502,359 participants from 22 assessment centers distributed over the United Kingdom. Most participants have genetic (97 %) and general physical measurement data (93 %) like the Framingham Score features available, while the more complex measures like metabolomics, retinal imaging or proteomics data are only available for a much smaller subset. (B) Ehrapy’s quality control workflow shows a distinct cluster of individuals (bottom right) from the Birmingham assessment center in a UMAP embedding of the retinal imaging data, which we further investigated and found to be artifacts of the image acquisition process, and thus ultimately excluded from subsequent analyses. (C) Myocardial Infarctions are recorded for 15 % of the male, and 7 % of the female study population. (D) For every modality and their respective combinations a linear Cox-Proportional hazards model was fitted to determine the prognostic potential of these for the myocardial Infarction phenotype. Commonly used cardiovascular risk factors show expected positive log-hazard ratios for increased blood pressure or total cholesterol, and negative ones for sampling age and female sex. (E) The predictive performance of individual modalities (dark blue) varies from a C-index of 0.63 to 0.77 for genetic and Framingham Risk respectively. Combining all features yields a C-index of 0.81.

Predictive performance for each modality was assessed by fitting Cox proportional hazards (**Figure 6C**) models on each of the feature sets using ehrapy (**Figure 6D**). The age of the first occurrence served as the time-to-event; alternatively, date of death or date of the last record in EHR served as censoring times. Models were evaluated using the Concordance Index (C-Index). Here, the combination of multiple modalities successfully improved the predictive performance for coronary heart disease by increasing the C-index from 0.63 (genetic), to 0.76 (genetics, age and sex) and to 0.77 (clinical predictors) with 0.81 (imaging and clinical predictors) for combinations of feature sets (**Figure 6E**). Our finding is in line with previous observations of complementary effects between different modalities, where a broader ‘major adverse cardiac event’ phenotype was modeled in the UKB with a neural network achieving a C-index of 0.72 using metabolomics, age and sex^77^. By analyzing the predictive potential of diverse data modalities alone and in combination through the same ehrapy analysis pipeline, we directly assessed the overlap of information contained in either one of them. We observe that adding genetic data improves the predictive potential in all cases because it is the only modality independent of the sampling age and only predicts the other modalities to a limited extent^78^. We also demonstrate that the addition of metabolomic data to the combination of clinical predictors, genetic information and imaging-derived measures did not improve predictive power (**Figure 6E**).

### Imaging-based disease severity projection through fate mapping

To demonstrate ehrapy’s ability to handle diverse image data and recover disease stages, we embedded pulmonary imaging data obtained from COVID-19 patients into a lower dimensional space and computationally inferred disease progression trajectories using pseudotemporal ordering. This describes a continuous trajectory or ordering of individual points based on feature similarity^79^, and has been extensively used e.g. in cellular analyses to recover opaque trajectories or lineages^57^. Continuous trajectories enable the map the fate of new patients onto precise states to potentially predict their future condition.

In COVID-19, a highly contagious respiratory illness caused by the coronavirus SARS-CoV-2 with symptoms ranging from mild flu-like symptoms to severe respiratory distress, chest X-ray images were described to exhibit opacities (bilateral patchy, ground glass) and consolidation in association with disease severity^80^.

We used COVID-19 chest X-ray images from the BrixIA^81^ dataset consisting of 192 images (**Figure 7A**) with expert annotations of disease severity. We made use of the scoring procedure of the BrixIA database which is based on six regions that were annotated by radiologists for disease severity ultimately resulting in a global score for disease severity classification (**Methods**). We embedded raw image features using a pre-trained DenseNet model (**Methods**) and further processed this embedding into a nearest-neighbors based UMAP space by the use of ehrapy (**Figure 7A**, **Methods**). Fate mapping based on imaging information (**Methods**) determined a severity ordering from mild to critical cases (**Figure 7B-D**). Images labeled as “normal” are projected to stay within the healthy group illustrating the robustness of our approach. Images of diseased patients were ordered by disease severity highlighting clear trajectories from “normal” to “critical” states despite the heterogeneity of the X-ray images stemming from e.g., different zoom (**Figure 7A**). To validate this raw image feature-based approach, we applied a similar workflow (**Methods**) to 790 eye retina images of patients affected by diabetic retinopathy and confirmed ehrapy’s potential to recover disease severity trajectories while differentiating severe and proliferative cases (**Supplementary Figure 8**). Here, we are able to additionally separate “Severe” from “Proliferate” cases into distinct clusters with unique trajectories.

**Figure 7.**
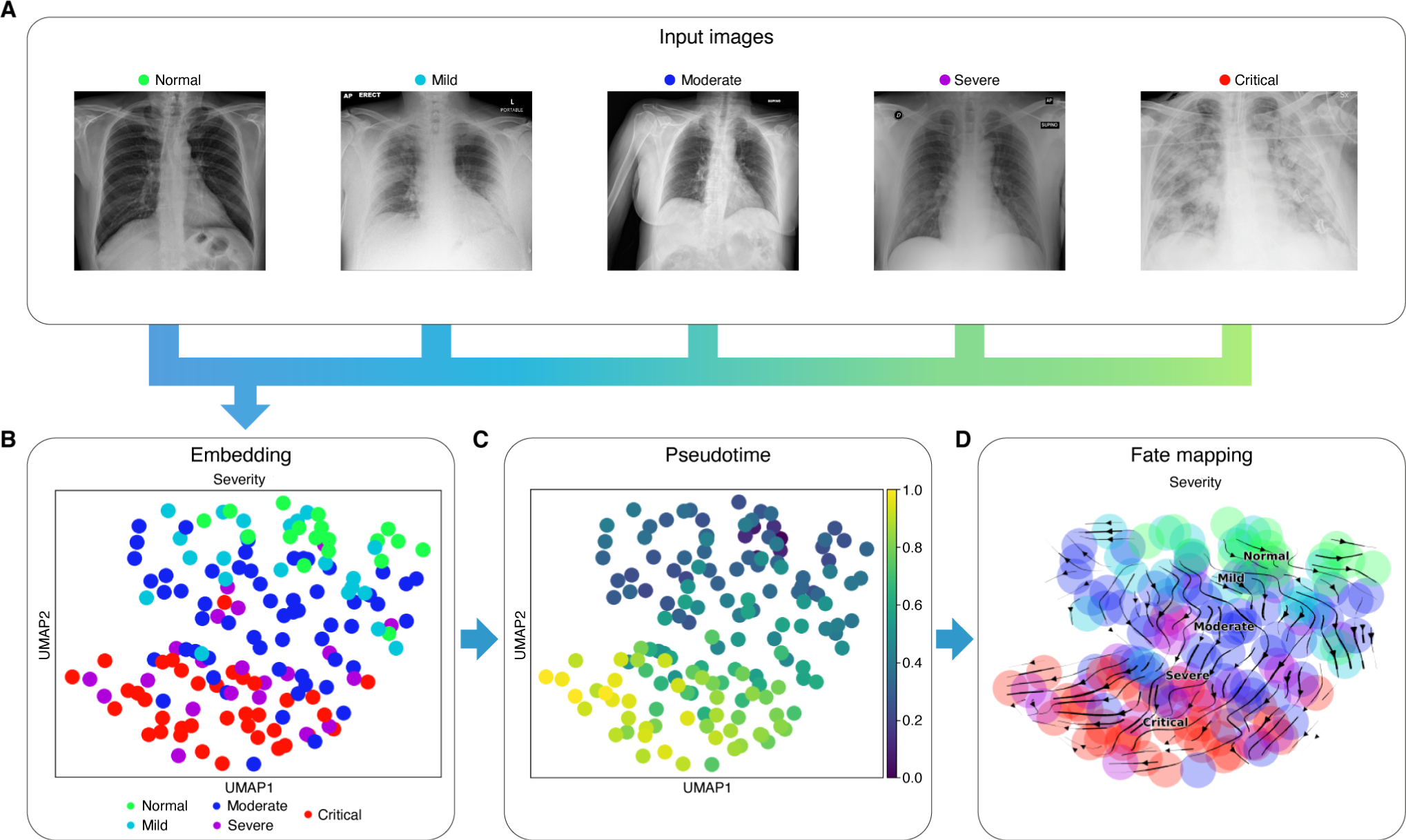
Recovery of disease severity trajectory in COVID 19 chest x-ray images. (A) Randomly selected example chest X-ray images of the BrixIA dataset highlight variance of BrixIA images. To determine fates, raw images are used as input for machine learning models that project them into a meaningful lower dimensional space. Using this representation, ehrapy determines pseudotime. Both, a nearest-neighbor graph and the determined pseudotime serve as input for the calculation of a transition matrix to enforce directionality. (B) UMAP visualization of the BrixIA dataset embedding shows a clear separation of disease classes. Disease severity was annotated by a senior radiologist of the BrixIA COVID-19 Project. (C) Calculated pseudotime for all images increases with distance to the “Normal” images. (D) Stream projection of fate mapping in UMAP space showcases disease severity trajectory of the COVID19 chest x-ray images.

## Discussion

By developing ehrapy’s structured data preparation, processing, and knowledge inference pipeline (**Figure 1**), we enable stratification of patient cohorts even in settings of highly variable data quality.

While using raw data directly acquired from clinical systems in conjunction with diagnostic labels from the PIC dataset, ehrapy successfully overcame limitations imposed by data missingness or oversimplification through diagnostic grouping or scoring processes. Ehrapy allowed us to identify fine-grained groups of “unspecified pneumonia” cases while discovering biomarkers and quantifying effects of medications on length of stay. Such retroactive characterization showcases ehrapy’s ability to generate novel perspectives or put complex evidence into context. Ultimately, such approaches can support feedback loops to improve diagnostic and therapeutic strategies leading to more efficiently allocated resources in healthcare.

Moreover, ehrapy’s flexible data structures enabled us to integrate the heterogeneous UKB data for predictive performance in myocardial infarction. The different data types and distributions posed a challenge for predictive models that were overcome with ehrapy’s preprocessing modules. Our analysis underscores the potential of integrating diverse modalities through ehrapy to enhance risk prediction by combined analysis of phenotypic and health outcome data at population scale.

By adapting pseudotime approaches that are commonly used in other omics domains, we successfully recovered disease trajectories from raw imaging data with ehrapy. While the diversity of the images made it challenging to find a suitable lower dimensional representation, ehrapy’s support for deep learning allowed us to quickly iterate on our results. The determined pseudotime, however, only orders data but does not necessarily provide a future projection per patient. Understanding the driver features for fate mapping in image-based data sets is challenging. The incorporation of image segmentation approaches could mitigate this issue and provide a deeper insight into the spatial and temporal dynamics of disease-related processes. Our approach goes beyond individual assessment including rater-based scoring to identify global trends, saving time while improving objectiveness.

Analysis limitations include the lack of control for informative missingness where the absence of information represents information in itself^82^. Translation such as in the PIC database from Chinese into English, results in potential information loss and inaccuracies as the Chinese ICD-10 code for diagnostics is an augmented version with seven characters compared to the five characters of the English version. Furthermore, legal restrictions or incompleteness of databases such as the lack of radiology images in the PIC database, cannot be accounted for. Further, low sample sizes or underrepresentation of e.g. non-white ancestries as well as biases participant self-selection limit ehrapy’s performance. This restricts deeper phenotyping of, for example, all “unspecified pneumonia” cases with respect to their survival which could be overcome by the use of multiple databases. Moreover, our causal inference use-case excludes the consideration of unrecorded variables such as SOFA scores for the causal graph. We counterbalanced this by employing several refutation methods that statistically reject the causal hypothesis such as a placebo treatment, a random common cause, or an unobserved common cause.

These diverse use-cases illustrate ehrapy’s potential to sufficiently address the need for a computationally efficient, extendable, reproducible, and easy-to-use framework. More specifically, ehrapy manages the lack of standardization regarding the collection of data as well as their on-disk storage formats through its compatibility with major standards such as Observational Medical Outcomes Partnership (OMOP), Common Data Model (CDM)^39^, HL7 or openEHR with extensive support for common tabular data formats. Subsequent sharing of analysis results is made easy through AnnData objects that can be stored and read independent of the used platform. As ehrapy is scalable to datasets that are much larger in size, it further supports analyses combining multiple data sources through its latent space integration algorithms, thereby enabling across-age, across-ethnicity and across-clinical or -study analysis, driving evidence in health care. Ehrapy’s rich documentation of the application programming interface (API) and extensive hands-on tutorials make EHR analysis accessible to both novices and experienced analysts.

As ehrapy remains under active development, users can expect ehrapy to continuously evolve, adding more efficient and flexible data representations. Through a tighter integration into the scverse ecosystem, we are planning to provide better support for the joint analysis of EHR, genetics and molecular data. Ehrapy has the potential to serve as a bridge between the EHR and the omics communities to expedite the development of methods for joint analysis. Building on top of ehrapy, we envision the development of EHR specific latent factor extraction^83^ and query-to-reference mapping methods to enable EHR atlas building^84^. To further promote the sharing and collective analysis of EHR data we envision adapted versions of interactive single-cell data explorers like cellxgene^85^, the UCSC cell browser^86^, or the Single Cell Expression Atlas^87^ for EHR data. Additional modules specifically for high frequency time-series data, natural language processing, and other data types are currently under development. The first community packages such as an ehrapy based version of TimeAx^88^ or clehrity (https://github.com/blengerich/clehrity) for EHR deconfounding are already available. Ehrapy together with a lively ecosystem of packages has the potential to enhance the scientific discovery pipeline to shape the era of EHR analysis.

## Methods

### Implementation of ehrapy

ehrapy has been implemented in the Python programming language and builds upon a number of existing numerical and scientific open-source libraries, specifically, matplotlib^89^, seaborn^90^, NumPy^47^, numba^91^, Scipy^92^, scikit-learn^45^, and Pandas^93^. All functions are grouped into task specific modules whose implementation is complemented with additional dependencies.

#### Data preparation

##### Dataloaders

Ehrapy is compatible with any type of vectorized data, where vectorized refers to the data being stored in structured tables in either on-disk or database form. The input and output module of ehrapy provides readers for common formats such as OMOP, csv tables, or SQL databases through Pandas. When reading in such datasets the data is stored in the appropriate slots in a new AnnData^38^ object. Ehrapy’s data module provides access to more than 20 public EHR datasets that feature diseases including, but not limited to, Parkinson’s disease, breast cancer, chronic kidney disease, and more. All dataloaders return AnnData objects to allow for immediate analysis.

##### AnnData for EHR data

The AnnData format serves as a pivotal structure in the realm of single-cell genomics. At its core, an AnnData object encapsulates diverse components, providing a holistic representation of data and metadata that are always aligned in dimensions and easily accessible. A Data Matrix (commonly referred to as “X”) stands as the foundational element, embodying the measured data. This matrix can be either dense, sparse, or ragged where dimensions do not align within the data matrix. The AnnData object can feature several such data matrices stored in “layers”. Examples of such layers can be unnormalized or unencoded data. These data matrices are complemented by an observations (commonly referred to as “obs”) segment where annotations on the level of patients or visits are stored. Patients’ age or sex, for instance, are often utilized as such annotations. The Variables (commonly referred to as “var”) section complements the observations, offering supplementary details about the features in the dataset such as missing data rates. The observation-specific matrices (commonly referred to as “obsm”) section extends the capabilities of the AnnData structure by allowing the incorporation of observation-specific matrices. These matrices can represent various types of information at the individual cell level, such as principal component analysis (PCA) results, t-distributed stochastic neighbor embedding (t-SNE) coordinates, or other dimensionality reduction outputs. Analogously, AnnData features a variables-specific variables (commonly referred to as “varm”) component. The observation-specific pairwise relationships (commonly referred to as “obsp”) segment complements the “obsm” section by accommodating observation-specific pairwise relationships. This can include connectivity matrices, indicating relationships between patients. The inclusion of an unstructured annotations (commonly referred to as “uns”) component further enhances flexibility. This segment accommodates unstructured annotations or arbitrary data that might not conform to the structured observations or variables categories.

Any AnnData object can be stored on disk in h5ad or Zarr format to facilitate data exchange.

##### Feature annotation

After AnnData creation, any metadata can be mapped against ontologies using Bionty (https://github.com/laminlabs/bionty). Bionty provides access to the Human Phenotype, Phecodes, Phenotype and Trait, Drug, Mondo, and Human Disease ontologies.

Key medical terms stored in an AnnData object in free text can be extracted using the Medical Concept Annotation Toolkit (medcat)^94^.

#### Data processing

##### Basic preprocessing and quality control

Ehrapy encompasses a suite of functionalities for fundamental data processing that are adopted from scanpy^44^ but adapted to EHR data:

1. **Regress out:** To address unwanted sources of variation, a regression procedure is integrated, enhancing the dataset’s robustness.
2. **Sumsample:** Select a specified fraction of observations.
3. **Highly variable features:** The identification and annotation of highly variable features following the “highly variable genes” function of scanpy is seamlessly incorporated, providing users with insights into pivotal elements influencing the dataset.

To identify and minimize quality issues, ehrapy provides several quality control functions:

1. **Basic quality control:** Determines the relative and absolute number of missing values per feature and per patient.
2. **Winsorization:** For data refinement, ehrapy implements a Winsorization process, creating a version of the input array less susceptible to extreme values.
3. **Feature clipping:** Imposes limits on features to enhance dataset reliability.
4. **Summarize features:** Calculates statistical indicators per feature, including minimum, maximum, and average values. This can be especially useful to reduce complex data with multiple measurements per feature per patient into sets of columns with single values.

Imputation is crucial in data analysis to address missing values, ensuring the completeness of datasets which can be required for specific algorithms. The “ehrapy” preprocessing module offers a range of imputation techniques:

1. **Explicit Impute:** Replaces missing values, either in all columns or a user-specified subset, with a designated replacement value.
2. **Simple Impute:** Imputes missing values in numerical data using mean, median, or the most frequent value, contributing to a more complete dataset.
3. **KNN Impute:** Utilizes K-nearest neighbor imputation to fill in missing values in the input AnnData object, preserving local data patterns.
4. **SoftImpute:** Applies the SoftImpute method for imputing missing values, offering a flexible and effective strategy for enhancing data completeness.
5. **IterativeSVD Impute:** Imputes missing values in an AnnData object using the IterativeSVD algorithm, contributing to improved data quality.
6. **MatrixFactorization Impute:** Utilizes MatrixFactorization for imputing missing data, offering a sophisticated approach to handling missing values in diverse datasets.
7. **NuclearNormMinimization Impute:** Implements NuclearNormMinimization for imputing missing data, enhancing the quality of imputed values through matrix factorization.
8. **MissForest Impute:** Implements the MissForest strategy for imputing missing data, providing a robust approach for handling complex datasets.
9. **MICE Impute:** Applies the Multiple Imputation by Chained Equations (MICE) algorithm for imputing data. This implementation is based on the miceforest (https://github.com/AnotherSamWilson/miceforest) package.

Data encoding can be required if categoricals are a part of the dataset to obtain numerical values only. Most algorithms in ehrapy are only compatible with numerical values. Ehrapy offers a wide range of encoding algorithms based on scikit-learn^45^:

1. **One-Hot Encoding:** Transforms categorical variables into binary vectors, creating a binary feature for each category and capturing the presence or absence of each category in a concise representation.
2. **Label Encoding:** Assigns a unique numerical label to each category, facilitating the representation of categorical data as ordinal values and supporting algorithms that require numerical input.
3. **Count Encoding:** Replaces categorical values with their respective frequency counts, offering a robust encoding strategy that captures the prevalence of each category in the dataset.
4. **Hash Encoding:** Utilizes hashing functions to convert categorical values into numerical representations, providing a space-efficient encoding method suitable for high-cardinality categorical features.

To ensure that the distributions of the heterogeneous data are aligned, ehrapy offers several normalization procedures:

1. **Log Normalization:** Applies the natural logarithm function to the data, useful for handling skewed distributions and reducing the impact of outliers.
2. **Max-Abs Normalization:** Scales each feature by its maximum absolute value, ensuring that the maximum absolute value for each feature is 1.
3. **Min-Max Normalization:** Transforms the data to a specific range (commonly [0, 1]) by scaling each feature based on its minimum and maximum values.
4. **Power Transformation Normalization:** Applies a power transformation to make the data more Gaussian-like, often useful for stabilizing variance and improving the performance of models sensitive to distributional assumptions.
5. **Quantile Normalization:** Aligns the distributions of multiple variables, ensuring that their quantiles match, which can be beneficial for comparing datasets or removing batch effects.
6. **Robust Scaling Normalization:** Scales data using the interquartile range (IQR), making it robust to outliers and suitable for datasets with extreme values.
7. **Scaling Normalization:** Standardizes data by subtracting the mean and dividing by the standard deviation, creating a distribution with a mean of 0 and a standard deviation of 1.
8. **Square Root Normalization:** Applies the square root function to the data, useful for reducing the impact of outliers and handling right-skewed distributions.
9. **Offset to Positive Values:** Shifts all values by a constant offset to make all values non-negative, with the lowest negative value becoming 0.

Dataset shifts can be corrected using the scanpy implementation of the combat^95^ algorithm, which employs a parametric and non-parametric empirical Bayes framework for adjusting data for batch effects that is robust to outliers.

Finally, a neighbors graph can be efficiently computed using scanpy’s implementation.

##### Embeddings

To obtain meaningful lower-dimensional embeddings that can subsequently be visualized and reused for downstream algorithms, ehrapy provides the following algorithms based on scanpy’s implementation:

1. **t-SNE (t-distributed Stochastic Neighbor Embedding):** Utilizes a probabilistic approach to embed high-dimensional data into a lower-dimensional space, emphasizing the preservation of local similarities and revealing clusters in the data.
2. **UMAP (Uniform Manifold Approximation and Projection):** Embeds data points by modeling their local neighborhood relationships, offering an efficient and scalable technique that captures both global and local structures in high-dimensional data.
3. **Force-Directed Graph Drawing:** Utilizes a physical simulation to position nodes in a graph, with edges representing pairwise relationships, creating a visually meaningful representation that emphasizes connectedness and clustering in the data.
4. **Diffusion Maps:** Applies spectral methods to capture the intrinsic geometry of high-dimensional data by modeling diffusion processes, providing a way to uncover underlying structures and patterns.
5. **Density Calculation in Embedding:** Quantifies the density of observations within an embedding, considering conditions or groups, offering insights into the concentration of data points in different regions and aiding in the identification of densely populated areas.

##### Clustering

Ehrapy further provides algorithms for clustering and trajectory inference based on scanpy:

1. **Leiden Clustering:** Utilizes the Leiden algorithm to cluster observations into subgroups, revealing distinct communities within the dataset with an emphasis on intra-cluster cohesion.
2. **Hierarchical Clustering Dendrogram:** Constructs a dendrogram through hierarchical clustering based on specified groupby categories, illustrating the hierarchical relationships among observations and facilitating the exploration of structured patterns.

##### Group comparisons

To compare any obtained clusters to obtain marker features that are significantly different between the groups, ehrapy extends scanpy’s “rank genes groups”. The original implementation which features a T-test for numerical data is complemented by a G-test for categorical data.

##### Dataset integration

Based on scanpy’s “ingest” function, ehrapy facilitates the integration of labels and embeddings from a well-annotated reference dataset into a new dataset, enabling the mapping of cluster annotations and spatial relationships for consistent comparative analysis. This process ensures harmonized clinical interpretations across datasets, especially useful when dealing with multiple experimental diseases or batches.

#### Knowledge inference

##### Survival analysis

Ehrapy’s implementation of survival analysis algorithms is based on lifelines (https://github.com/CamDavidsonPilon/lifelines):

1. **Ordinary Least Squares (OLS) Model:** Creates a linear regression model using Ordinary Least Squares (OLS) from a specified formula and an AnnData object, allowing for the analysis of relationships between variables and observations.
2. **Generalized Linear Model (GLM):** Constructs a Generalized Linear Model (GLM) from a given formula, distribution, and AnnData, providing a versatile framework for modeling relationships with non-linear data structures.
3. **Kaplan-Meier:** Fits the Kaplan-Meier estimate to generate survival curves, offering a visual representation of the probability of survival over time in a dataset.
4. **Cox Hazard Model:** Constructs a Cox Proportional-Hazards model using a specified formula and an AnnData object, enabling the analysis of survival data by modeling the hazard rates and their relationship to predictor variables.
5. **Logrank Test:** Calculates the p-value for the logrank test, comparing the survival functions of two groups, providing statistical significance for differences in survival distributions.
6. **GLM comparison:** Given two fitted Generalized Linear Models (GLMs), where the larger encompasses the parameter space of the smaller, this function returns the p-value indicating the significance of the larger model adding explanatory power beyond the smaller model.

##### Trajectory inference

Trajectory inference is a computational approach that reconstructs and models the developmental paths and transitions within heterogeneous clinical data, providing insights into the temporal progression underlying complex systems. Ehrapy offers several inbuilt algorithms for trajectory inference based on scanpy:

1. **Diffusion Pseudotime:** Infers the progression of observations by measuring geodesic distance along the graph, providing a pseudotime metric that represents the developmental trajectory within the dataset.
2. **PAGA (Partition-based Graph Abstraction):** Maps out the coarse-grained connectivity structures of complex manifolds using a partition-based approach, offering a comprehensive visualization of relationships in high-dimensional data and aiding in the identification of macroscopic connectivity patterns.

Since ehrapy is compatible with scverse, further trajectory inference based algorithms such as cell rank can be seamlessly applied.

##### Causal Inference

Ehrapy’s causal inference module is based on dowhy^68^. It is based on four key steps that are all implemented in ehrapy:

1. **Graphical Model Specification:** Define a causal graphical model representing relationships between variables and potential causal effects.
2. **Causal Effect Identification:** Automatically identify whether a causal effect can be inferred from the given data, addressing confounding and selection bias.
3. **Causal Effect Estimation:** Employ automated tools to estimate causal effects, utilizing methods like matching, instrumental variables, or regression.
4. **Sensitivity Analysis and Testing:** Perform sensitivity analysis to assess the robustness of causal inferences and conduct statistical testing to determine the significance of the estimated causal effects.

##### Patient stratification

Ehrapy’s complete pipeline from preprocessing to the generation of lower dimensional embeddings, clustering, statistical comparison between determined groups, and more facilitates the stratification of patients.

#### Visualization

Ehrapy features an extensive visualization pipeline that is customizable and yet offers reasonable defaults. Almost every analysis function is matched with at least one visualization function that often shares the name but is available through the plotting module. For example, after importing ehrapy as “ep”, “ep.tl.umap(adata)” runs the UMAP algorithm on an AnnData object, while “ep.pl.umap(adata)” would then plot a scatterplot of the UMAP embedding.

Ehrapy further offers a suite of more generally usable and modifiable plots:

1. **Scatter Plot:** Visualizes data points along observation or variable axes, offering insights into the distribution and relationships between individual data points.
2. **Heatmap:** Represents feature values in a grid, providing a comprehensive overview of the data’s structure and patterns.
3. **Dot Plot:** Displays count values of specified variables as dots, offering a clear depiction of the distribution of counts for each variable.
4. **Filled Line Plot:** Illustrates trends in data with filled lines, emphasizing variations in values over a specified axis.
5. **Violin Plot:** Presents the distribution of data through mirrored density plots, offering a concise view of the data’s spread.
6. **Stacked Violin Plots:** Combines multiple violin plots, stacked to allow for visual comparison of distributions across categories.
7. **Group Mean Heatmap:** Creates a heatmap displaying the mean count per group for each specified variable, providing insights into group-wise trends.
8. **Hierarchically-Clustered Heatmap:** Utilizes hierarchical clustering to arrange data in a heatmap, revealing relationships and patterns among variables and observations.
9. **Rankings Plot:** Visualizes rankings within the data, offering a clear representation of the order and magnitude of values.
10. **Dendrogram Plot:** Plots a dendrogram of categories defined in a groupby operation, illustrating hierarchical relationships within the dataset.

The implementation of ehrapy is accessible at https://github.com/theislab/ehrapy together with extensive API documentation at https://ehrapy.org.

### Paediatric Intensive Care database analysis

#### Study design

We collected clinical data from the Paediatric Intensive Care (PIC)^33^ version 1.1.0 database. PIC is a single-center, bilingual (English and Chinese) database hosting information of children admitted to critical care units at the Children’s Hospital of Zhejiang University School of Medicine in China. The requirement for individual patient consent was waived since the study did not impact clinical care, and all protected health information was de-identified. The database contains 13,499 distinct hospital admissions of 12,881 distinct paediatric patients. These were admitted to 5 intensive critical care units (ICUs) with 119 total critical care beds: general ICU, paediatric ICU (PICU), surgical ICU (SICU), cardiac ICU (CICU), and neonatal ICU (NICU) between 2010 and 2018. The mean age of the patients is 2.5 years of which 42.5% were female. The in-hospital mortality was 7.1%, the mean hospital stay 17.6 days and the mean ICU stay 9.3 days. 468 (3.6%) of the patients were admitted multiple times. Demographics, diagnoses, doctors’ notes, laboratory and microbiology tests, prescriptions, fluid balances, vital signs, and radiographics reports were collected from all patients. For more details we refer to the original publication of Zeng et al^33^.

#### Study participants

Individuals aged > 18 years were excluded from the study. We grouped the data into 3 distinct groups: “Neonates” (0 to 28 days of age; 2968 patients), “infants” (1 month to 12 months of age; 4876 patients), and “youths” (13 months to 18 years of age; 6097 patients). We primarily analyzed the “youths” group with the discharge diagnosis ‘unspecified pneumonia’ (277 patients).

#### Data collection

The collected clinical data included demographics, laboratory and vital sign measurements, diagnoses, microbiology and medication information, and mortality outcomes. The 5-character 10th English International Statistical Classification of Diseases and Related Health Problems (ICD10) code was used whose values are based on the 7-character Chinese ICD-10 code.

#### Dataset extraction and analysis

We downloaded the PIC database of version 1.1.0 from Physionet^1^ to obtain 17 CSV tables. Using Pandas we selected all information with more than 50% coverage rate including demographics, laboratory and vital sign measurements (**Figure 2**). To reduce the amount of noise we only added the minimum, maximum and average of all measurements into the data matrix of the generated AnnData object. Exam reports were removed since they only describe diagnostics but not detailed findings. All further diagnoses, microbiology and medication information were included into the observations slot to ensure that the data was not used for the calculation of embeddings but still available for the analysis. This ensured that the embedding was not divided into treated and untreated groups but rather solely based on phenotypic features. We imputed all missing data through k-nearest neighbors imputation (k=20), log normalized and winsorized the data using ehrapy to obtain 261 ICU visits with 572 features. Of those 572 features, 254 were stored in the matrix X and the remaining 318 in the obs slot. We observed several features such as medications to distort UMAP embeddings into groups such as “received any medication” or “did not receive any medication” which wasn’t meaningful for our questions at hand. For clustering and visualization purposes we calculated 50 principal components to calculate a nearest neighbors graph to serve as input for Uniform Manifold Approximation and Projection (UMAP) on the AnnData object using ehrapy.

#### Patient stratification

We applied the community detection algorithm Leiden with resolution 0.6 on the nearest neighbor graph using ehrapy. The four obtained clusters served as input for two-sided t-tests for all numerical values and two-sided g-tests for all categorical values for all four clusters against the union of all three other clusters respectively. p-values were corrected with Benjamini-Hochberg^96^ procedure. Clusters were annotated by expert medical doctors based on the determined overrepresented features.

#### Kaplan-Meier survival analysis

Patients with up to 360 hours of total stay were selected for Kaplan-Meier survival analysis. Kaplan-Meier analysis was conducted with ehrapy between all four determined pneumonia groups. Significance was tested using a multivariate logrank test. An additional Kaplan-Meier analysis was conducted for all kids jointly concerning the liver markers AST, ALT, GGT. To determine whether measurements were inside or outside the norm range we used the following cutoffs:

Statistical significance was determined using a log rank test. P-values less than 0.05 were labeled significant.

#### Causal effect of mechanism of action on length of stay

##### Generation of administered medication categories

Medication names as specified in the PIC dataset with application forms removed were fuzzy-matched against all of drugbank 5.1^97^. The most likely matches were manually verified and their medication categories extracted leading to about 800 medication categories. Of these 800 categories we extracted corticosteroids, cephalosporins, antiviral agents, antifungal agents, carbapenems, and penicillins.

##### Causal Inference

Together with medical experts, we manually curated a minimal causal graph reflecting the interaction between the most important blood markers of pneumonia on length of stay and all applied interventions (here medication categories). Causal inference was conducted with ehrapy’s dowhy based causal inference module using the determined causal graph. All medication groups were treated as causal interventions (treatments) and length of stay was set as target variable (outcome). Linear regression was used as the estimation method. 4 patients with a length of stay of more than 90 days were excluded due to excessive length of stay. The refuters ‘placebo_treatment_refuter’, ‘random_common_cause’, ‘data_subset_refuter’, and ‘add_unobserved_common_cause’ were employed.

### UK Biobank analysis

#### Study population

We utilize information from the UK Biobank cohort, which represents a cross-section of the general UK population. This study involved the enrollment of individuals between 2006 and 2010 across 22 different assessment centers throughout the United Kingdom. The tracking of participants is still ongoing. Within the UK Biobank dataset, there is metabolomics data measured using NMR techniques available for a subset of individuals. Participants visited the assessment center up to four times and completed additional online followup questionnaires. Here, we restrict the analyses to data obtained from the initial assessment including the blood draw for obtaining the metabolomics data, and the retinal imaging as well as physical measures. This restricts the study population to 32,436 individuals for which all of these modalities are available. We have a clear study start point for each individual with the date of their initial assessment center visit. The study population has a mean age of 57 years, is 54 % female, and is censored at age 69 on average, 4.7 % experience an incident myocardial Infarction, and 8.1 % have prevalent type 2 diabetes. The study population comes from 6 of the 22 assessment centers due to the retinal imaging only being performed at those.

#### Data Collection

For the endpoint definition we rely on the first occurrence data available in the UK-Biobank which compiles the first date each diagnosis was recorded for a participant in a hospital in ICD-10 nomenclature. Subsequently, we map this data to Phecodes, and focus on Phecode 404.1 for myocardial Infarction.

The Framingham risk score was developed on data from 8491 participants of the framingham heart study to assess general cardiovascular risk. It includes easily obtainable predictors and is therefore easily applicable in clinical practice, although newer and more specific risk scores exist and might be used more frequently. It includes age, sex, smoking behavior, blood pressure, total and LDL-cholesterol as well as information on insulin, antihypertensive and cholesterol lowering medications, all of which are routinely collected in the UK-Biobank.

The Metabolomics data used in this study was obtained using proton nuclear magnetic resonance spectroscopy, a low-cost method with relatively low batch effects. It covers established clinical predictors like albumin and cholesterol as well as a range of lipids, amino acids and carbohydrate-related metabolites.

The retinal optical coherence tomography derived features were returned by researchers to the UK-Biobank^74,75^. They used the available scans and determined the macular volume, macular thickness, retinal pigment epithelium thickness, disc diameter, cup to disk ratio across different regions, as well as the thickness between the inner nuclear layer and external limiting membrane, inner and outer photoreceptor segments, and the retinal pigment epithelium across different regions.

#### Data analysis

After exporting the data from the UK-Biobank all time points were transformed into participant age entries. Only participants without prevalent myocardial Infarction (relative to the first assessment center visit at which all data was collected) were included.

The data was preprocessed for retinal imaging and metabolomics subsets separately, to enable a clear analysis of missing data, and allow for the k-nearest neighbors-based imputation (k=20) of missing values when less than 10 % were missing for a given participant. The genetics and framingham analyses were available for almost every participant, and not imputed, instead individuals without them were dropped from the analyses. Since genetic risk modeling poses entirely different methodological and computational challenges, we applied a published polygenic risk score for coronary heart disease using 6.6 million variants^72^. This was computed using the plink2 score option on the imputed genotypes available in the UKB.

UMAP spaces were computed using default ehrapy parameters on the full feature sets. Kaplain-Meier curves and Cox-proportional hazards models were fitted using ehrapy’s survival analysis module and the lifelines package. Models were evaluated using the C-index^98^ as a metric. It can be seen as an extension of the common area under the receiver operator characteristic score to time-to-event datasets, in which events are not observed for every sample.

#### In depth quality control of retina derived features

A UMAP plot of the retina-derived features indicating the assessment centers shows a cluster of samples that lie somewhat outside the general population and mostly attended the Birmingham assessment center (**Figure 6B**). To further investigate this, we performed Leiden clustering of resolution 0.3 (**Supplementary Figure 7A**) and isolated this group in cluster 5. When comparing cluster 5 to the rest of the population in the retina derived feature space, we noticed that many individuals in cluster 5 showed overall retinal pigment epithelium (RPE) thickness measures substantially elevated over the rest of the population in both eyes (**Supplementary Figure 7B**), which is mostly a feature of this cluster as indicated by UMAP plots color coding RPE thickness (**Supplementary Figure 7D, E**). To investigate potential confounding, we computed ratios between cluster 5 and the rest of the population over the obs DataFrame containing the Framingham features, diabetes-related phecodes and genetic principle components. Out of the top and bottom 5 highest ratios observed, 6 are in genetic principle components which are commonly used to represent genetic ancestry in a continuous space (**Supplementary Figure 7C**). Additionally, diagnoses for Type 1 and 2 diabetes and anti-hypertensive use are enriched in cluster 5. Further investigating the ancestry, we computed log ratios for self-reported ancestries and absolute counts, which showed no robust enrichment and depletion effects.

A closer look at three quality control measures of the Imaging pipeline, we found cluster 5 to be an outlier in terms of either image quality (**Supplementary Figure 7E**), or minimum motion correlation (**Supplementary Figure 7F**) and the Inner Limiting Membrane (ILM) Indicator (**Supplementary Figure 7G**), all of which can be indicative of artifacts in image acquisition and downstream processing^99^. Subsequently, we exclude 301 individuals from cluster 5 from all analyses.

### COVID-19 Chest-x-ray fate determination

#### Dataset overview

We used the public BrixIA COVID-19 Dataset which contains 192 chest X-ray images annotated with Brixia-scores^81^. Hereby, 6 regions were annotated by a senior with more than 20 years of experience and a junior radiologist from 0-3 (disease severity). A global score was determined which is the sum of all of these regions and therefore ranges from 0-18 (S-Global). S-Global scores of 0 were classified as normal. Images that only had severity values up to 1s in all 6 regions were classified as mild. Images with severity values greater or equal to 2, but a S-Global score of less than 7 were classified as moderate. All images that contained at least one 3 any of its 6 regions with a S-Global score between 7 and 10 were classified as severe and all remaining images with S-Global scores greater than 10 with at least one 3 were labeled critical. The dataset and instructions to download the images can be found here: https://github.com/ieee8023/covid-chestxray-dataset

#### Dataset extraction and analysis

All images were first resized to 224×224. Afterwards, the images underwent a random affine transformation that involved rotation, translation, and scaling. The rotation angle was randomly selected from a range of −45 to 45 degrees. The images were also subject to horizontal and vertical translation, with the maximum translation being 15% of the image size in either direction. Additionally, the images were scaled by a factor ranging from 0.85 to 1.15. The purpose of applying these transformations was to enhance the dataset and introduce variations, ultimately improving the robustness and generalization of the model.

To generate embeddings we used a pre-trained DenseNet model with weights ‘densenet121-res224-all’ of TorchXRayVision^100^. A DenseNet is a convolutional neural network which makes use of dense connections between layers (Dense Blocks) where all layers (with matching feature-map sizes) directly connect with each other. To maintain a feed-forward nature, every layer in the DenseNet architecture receives supplementary inputs from all preceding layers and transmits its own feature-maps to all subsequent layers. The model was trained on the *nih-pc-chex-mimic_ch-google-openi-rsna* dataset^101^.

The generated latent embedding was subject to principal component analysis, nearest neighbors graph calculation and a UMAP embedding with three components that was finally visualized using ehrapy. To determine fates, we employed CellRank^58^ with the PseudotimeKernel. We randomly picked a root in the group of images that were labeled ‘Normal’. We computed the transition matrix with a soft threshold scheme and a projection on top of the UMAP embedding.

### Diabetic retinopathy fate determination

#### Dataset overview

We used a comprehensive dataset of 30646 high-resolution retina images of 15323 patients, captured under various imaging conditions, with paired left and right eye fields for each subject. Each image is associated with a subject ID and is classified by a clinician for the presence of diabetic retinopathy (DR) using a five-level severity scale ranging from 0 (no DR; normal) to 4 (proliferative DR). Notably, the dataset showcases variations in imaging models and camera types, leading to differing visual appearances between left and right eye images. Importantly, images may be anatomically accurate or presented in an inverted manner, often indicated by the relative positions of the macula and optic nerve or the presence of a notch.

#### Dataset extraction and analysis

All images were initially resized to a dimension of 256×256, followed by center cropping to a final size of 224×224. Subsequently, these images underwent random horizontal flipping with a 50% probability of being flipped. After this, an augmentation process was applied using the RandAugment^102^ automated data augmentation pipeline, employing the ‘rand-m9-mstd0.5’ policy. This pipeline involved selecting two augmentation operations from a predefined set and applying them sequentially to the images with a specified magnitude. Each augmentation operation had a 50% probability of being applied. The set of 15 distinct augmentations included operations such as autocontrast, histogram equalization, image inversion (negation), random image rotation within a range of −10 to 10 degrees, polarization, solarization, solarization addition, color balance adjustment, contrast adjustment, brightness adjustment, sharpness adjustment, image shearing in the x- and y-directions, and image translation in the x- and y-directions. Following the augmentation process, the images were normalized using mean and standard deviation values. It’s important to note that all augmentations were exclusively applied during the training phase to enhance model generalization and were omitted during inference.

To generate embeddings, convolutional neural networks, specifically ResNet18, were employed. The network was first fine-tuned to classify the diabetic retinopathy progression scale. The weights of the convolutional layers were initialized using the pretrained ResNet18 model on the ImageNet dataset. The fully connected layers from the original ResNet18 were replaced with a new set of fully connected layers designed to predict 5 classes of diabetic retinopathy progression scale. Given the significant class imbalance, a weighted random sampler was additionally utilized during training to ensure an equal number of examples were sampled from each class.

Once the model was trained, the output of the penultimate layer was used to visualize the alignment of samples in the activation space. Subsequently, the latent embeddings extracted from the fine-tuned network underwent PCA. Following PCA, a nearest neighbors graph was calculated, and a UMAP embedding was generated using ehrapy. To determine cell fates, CellRank was employed, utilizing the PseudotimeKernel and a ‘soft’ threshold scheme for transition matrix computation. For the determination of fates, a random image from the group of images labeled as ‘Normal’ was selected as a root. The computed transition matrix was then projected onto the UMAP embedding. Due to the class imbalance reasons we subsampled all images to 790 images to have 158 records per class for clearer visualizations.

## Code and data availability

The ehrapy source code is available at https://github.com/theislab/ehrapy under the Apache 2.0 license. Further documentation, tutorials and examples are available at https://ehrapy.org.

Jupyter notebooks to reproduce our analysis and figures including Conda environments that specify all versions are available at https://github.com/theislab/ehrapy_reproducibility.

Physionet provides access to the PIC database at https://physionet.org/content/picdb/1.1.0 for credentialed users. The BrixIA images are available at https://github.com/BrixIA/Brixia-score-COVID-19. The diabetic retinopathy dataset is available at https://www.kaggle.com/c/diabetic-retinopathy-detection/data. The data used in this study were obtained from the UK Biobank (www.ukbiobank.ac.uk). Access to the UK Biobank resource was granted under application number 49966. The data are available to researchers upon application to the UK Biobank in accordance with their data access policies and procedures.

## Acknowledgements

We thank Meshal Ansari who designed the ehrapy logo. The authors thank F. Alexander Wolf, Malte Lücken, Jakob Steinfeldt, Benjamin Wild, Gunnar Rätsch, and Dennis Shung for feedback on the project. We further thank Lennard Halle, Yuge Ji, Malte Lücken and Raphael Kfuri Rubens for constructive comments on the manuscript. This research was conducted using data from UK Biobank, a major biomedical database (https://www.ukbiobank.ac.uk) via application no. 49966.

## Author contributions

LH and FJT conceived the study. LH, PE, XZ, AN, LZ, VS, TT, LeH, NH, RK and IV implemented ehrapy. LH, PE, NL, LS, TT, and AH analyzed the PIC database. JUzB and LH analyzed the UK biobank database. XZ and LH analyzed the COVID-19 chest X-ray dataset. NH and LH analyzed the diabetic retinopathy dataset. LH, PE, and JUzB wrote the manuscript. FJT, AH, HBS, and RE supervised the work. All authors read, corrected and approved the final manuscript.

## Supplements

### Supplementary Figures

**Supplementary Figure 1:**
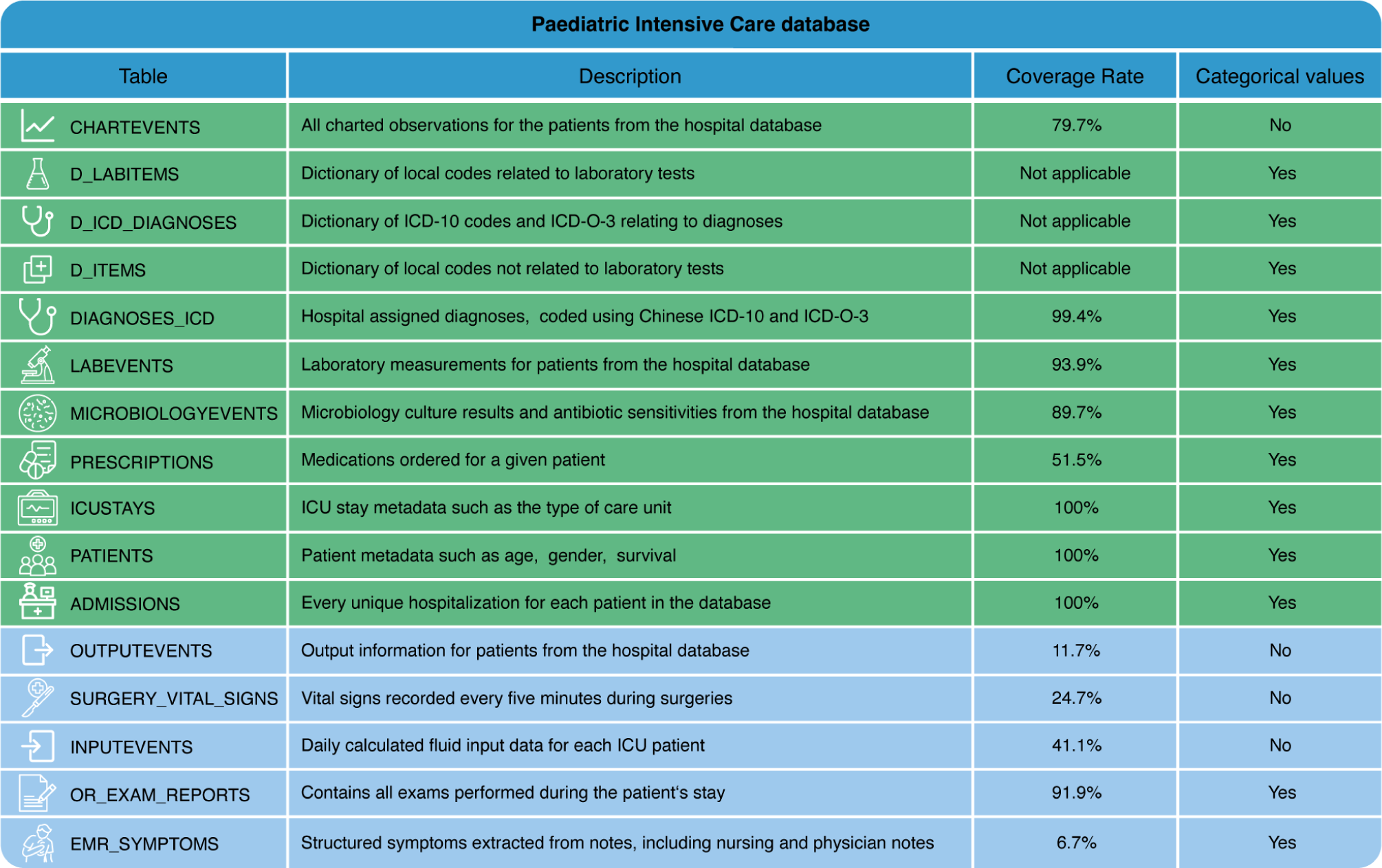
Overview of the paediatric intensive care database (PIC). The database consists of several tables corresponding to several data modalities and measurement types. All tables colored in green were selected for analysis and all tables in blue were discarded based on coverage rate. Despite the high coverage rate, we discarded the “OR_EXAM_REPORTS” table because of lack of detail in the exam reports.

**Supplementary Figure 2.**
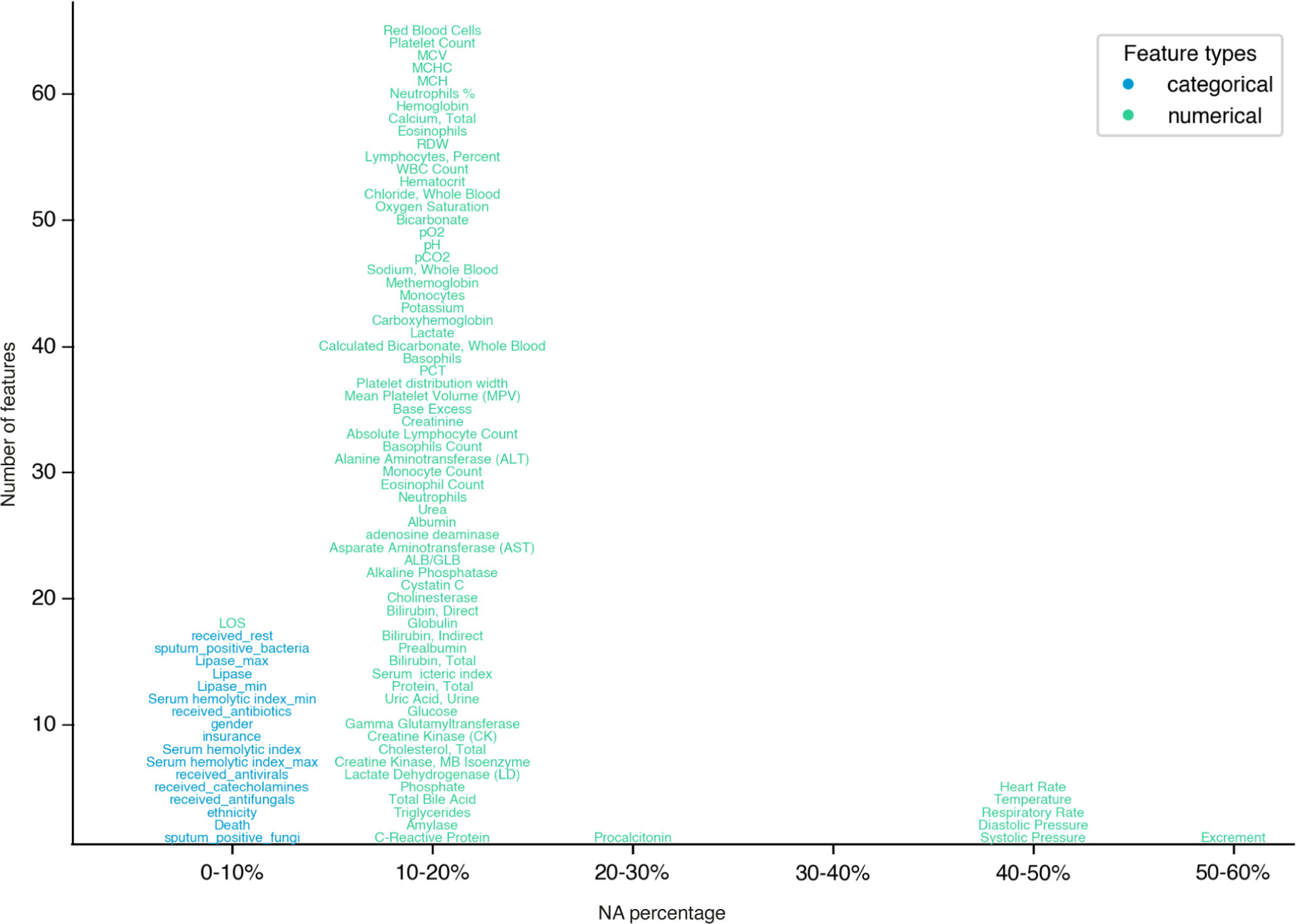
Missing data distribution for the “youths” group of the PIC dataset. The x-axis represents the percentage of missing values in each feature. The y-axis reflects the number of features in each bin with text labels representing the names of the individual features.

**Supplementary Figure 3.**
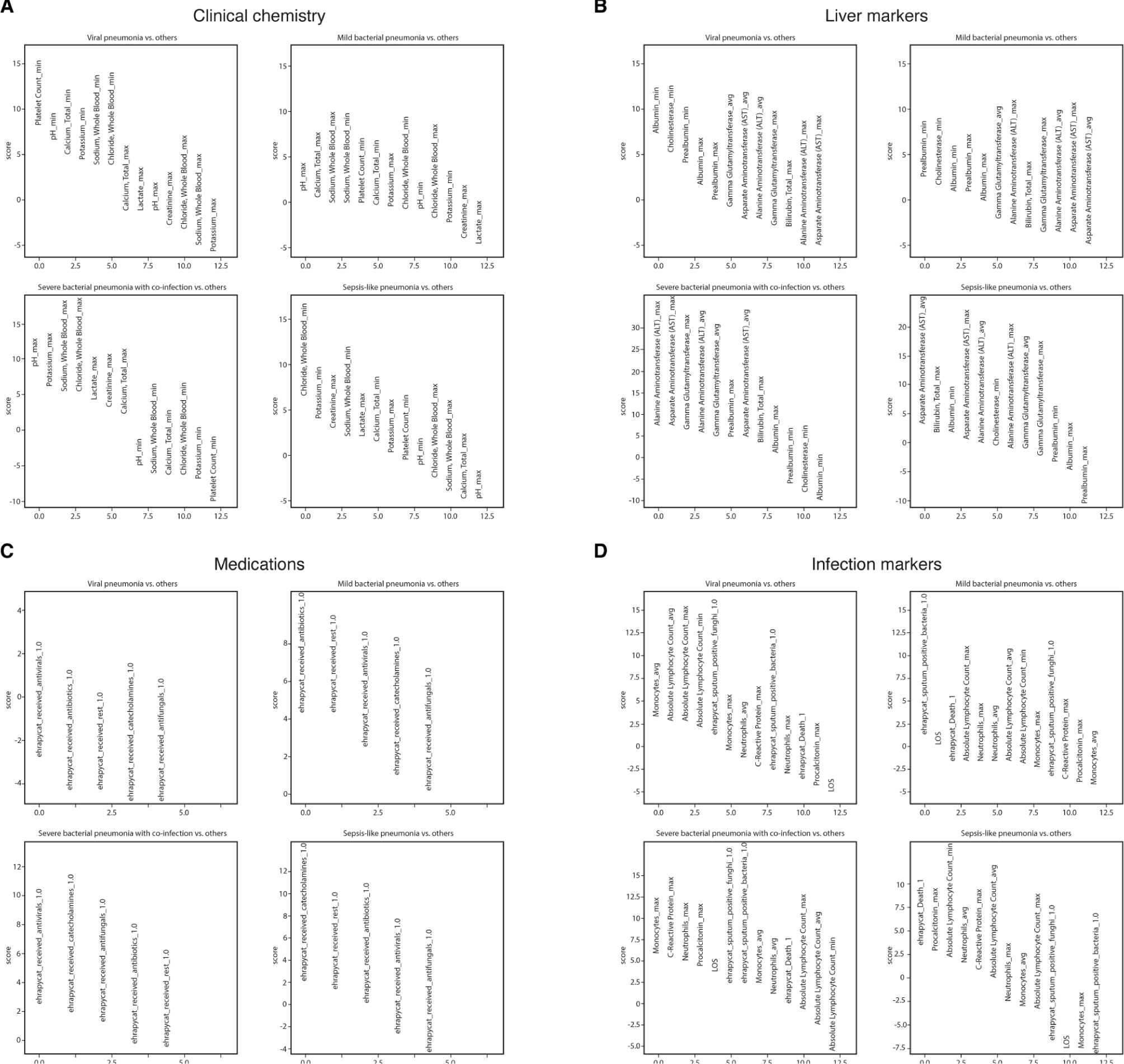
Feature rankings of stratified patient groups. Scores reflect the z-score underlying the p-value per measurement for each group. Higher scores (above 0) reflect overrepresentation of the measurement compared to all other groups and vice versa. (A) By clinical chemistry. (B) By liver markers. (C) By medication type. (D) By infection markers.

**Supplementary Figure 4.**
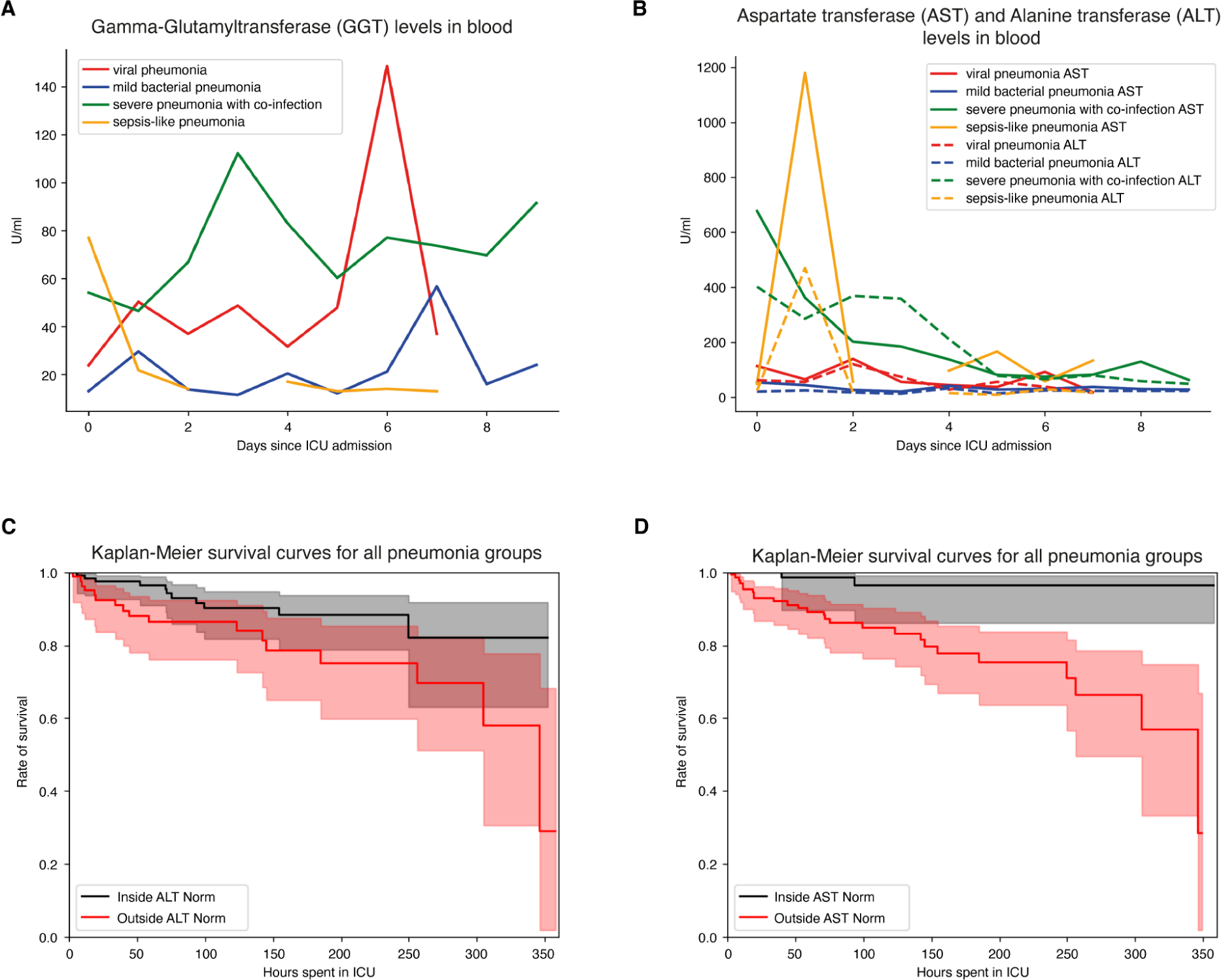
Liver marker value progression for the “youths” group and Kaplan-Meier curves. (A) Viral and severe pneumonia with co-infection groups display enriched gamma-glutamyl transferase levels in blood serum. (B) Aspartate transferase (AST) and Alanine transaminase (ALT) levels are enriched for severe pneumonia with co-infection during early ICU stay. (C) and (D) Kaplan Meier curve for ALT and AST demonstrate lower survivability for children with measurements outside the norm.

**Supplementary Figure 5.**
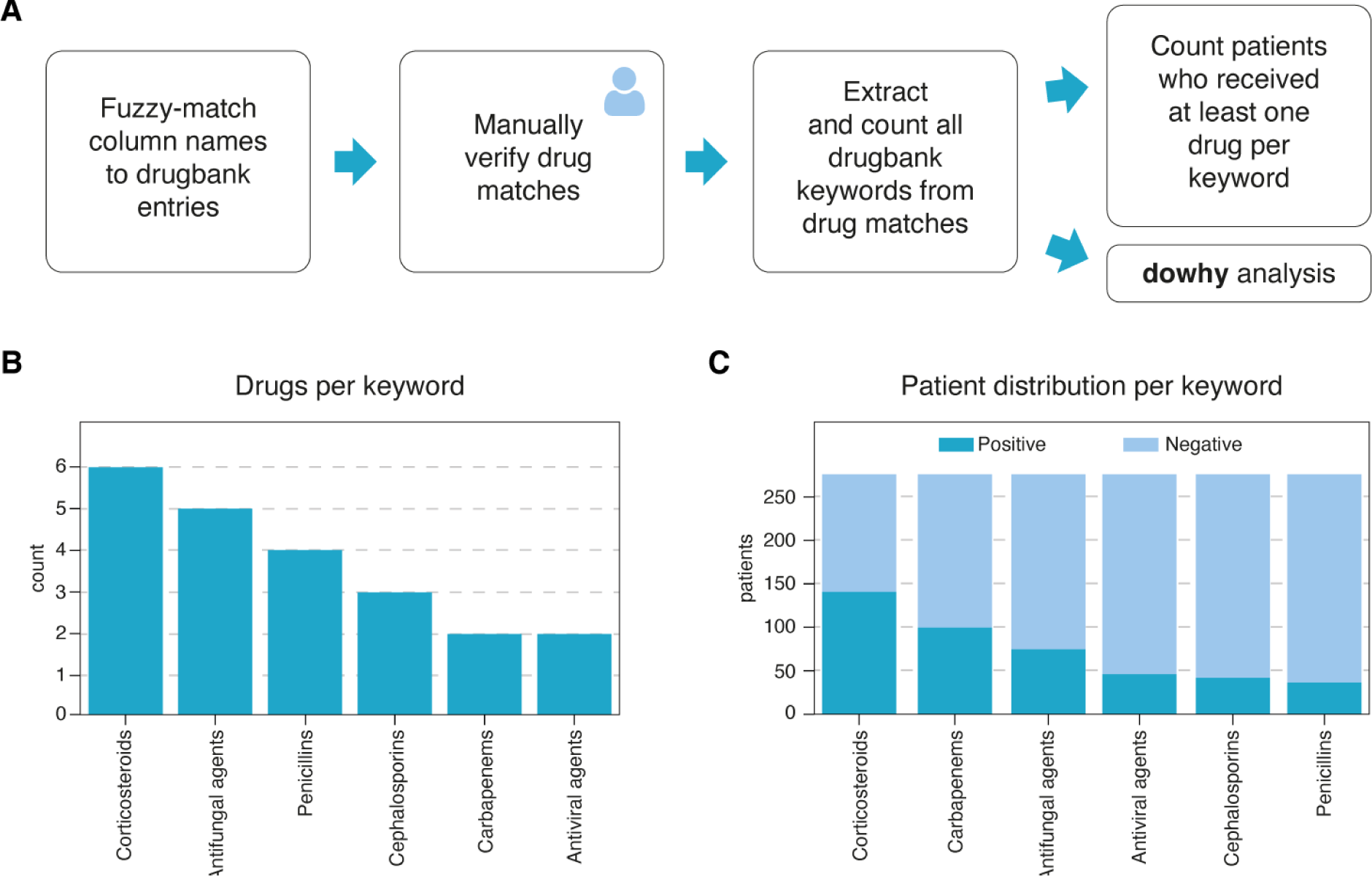
Overview of medication categories used for causal inference. (A) Feature engineering process to group administered medications into medication categories using drugbank. (B) Number of medications per medication category. (C) Number of patients that received (dark blue) and did not receive specific medication categories (light blue).

**Supplementary Figure 6.**
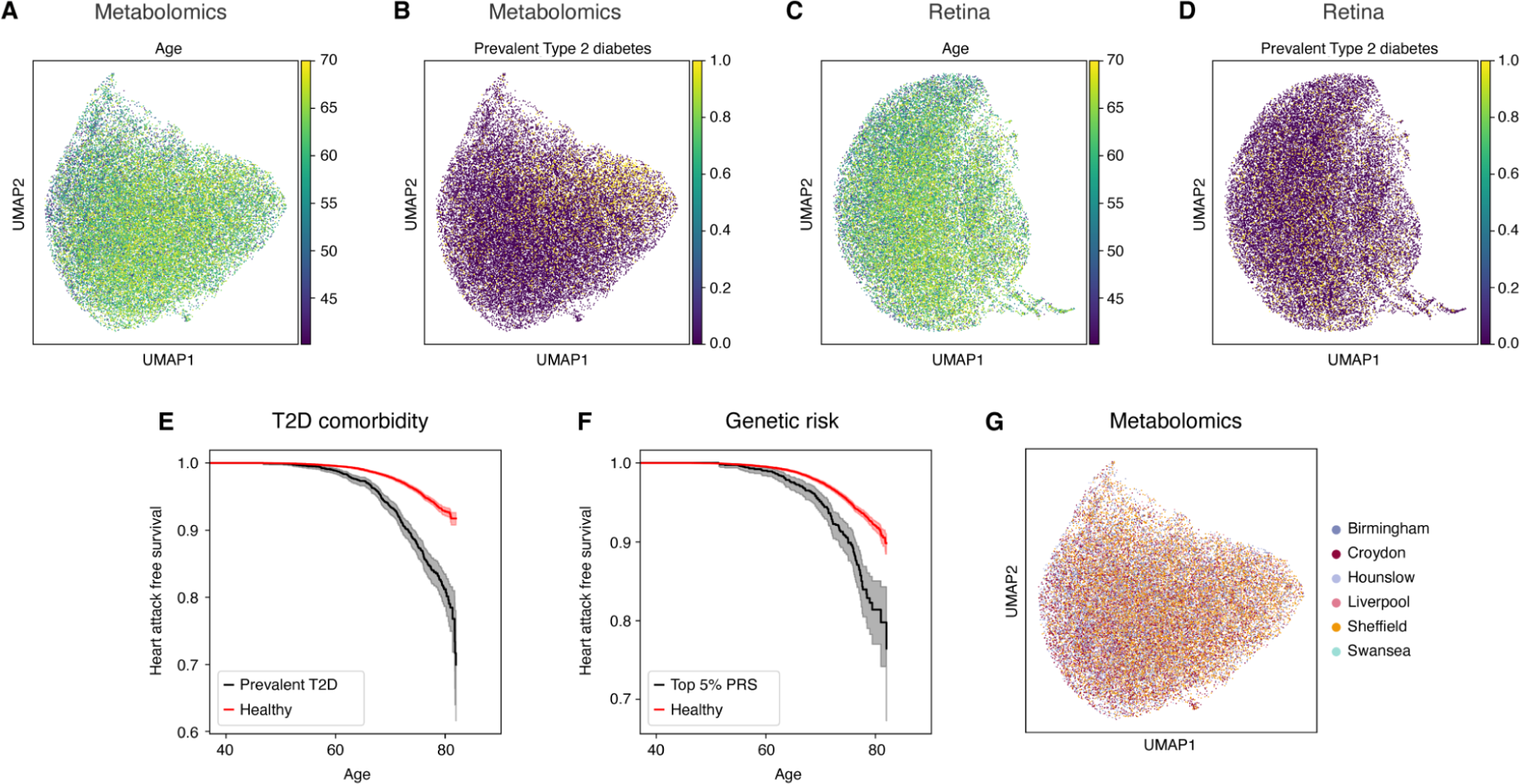
UK-Biobank data overview and quality control across modalities. (A) UMAP plot of the metabolomics demonstrating clear gradient with respect to age at sampling), and (B) type 2 diabetes prevalence. (C) Analogously, the features derived from retinal imaging show a less pronounced age gradient, and (D) type 2 diabetes prevalence gradient. (E) Stratifying myocardial Infarction risk by the type 2 diabetes comorbidity confirms vastly increased risk with a prior type 2 diabetes diagnosis. (F) Similarly, the polygenic risk score for coronary heart disease used in this work substantially enriches myocardial Infarction risk in its top 5% percentile. (G) UMAP visualization of the metabolomics features colored by the assessment center shows no discernable biases.

**Supplementary Figure 7.**
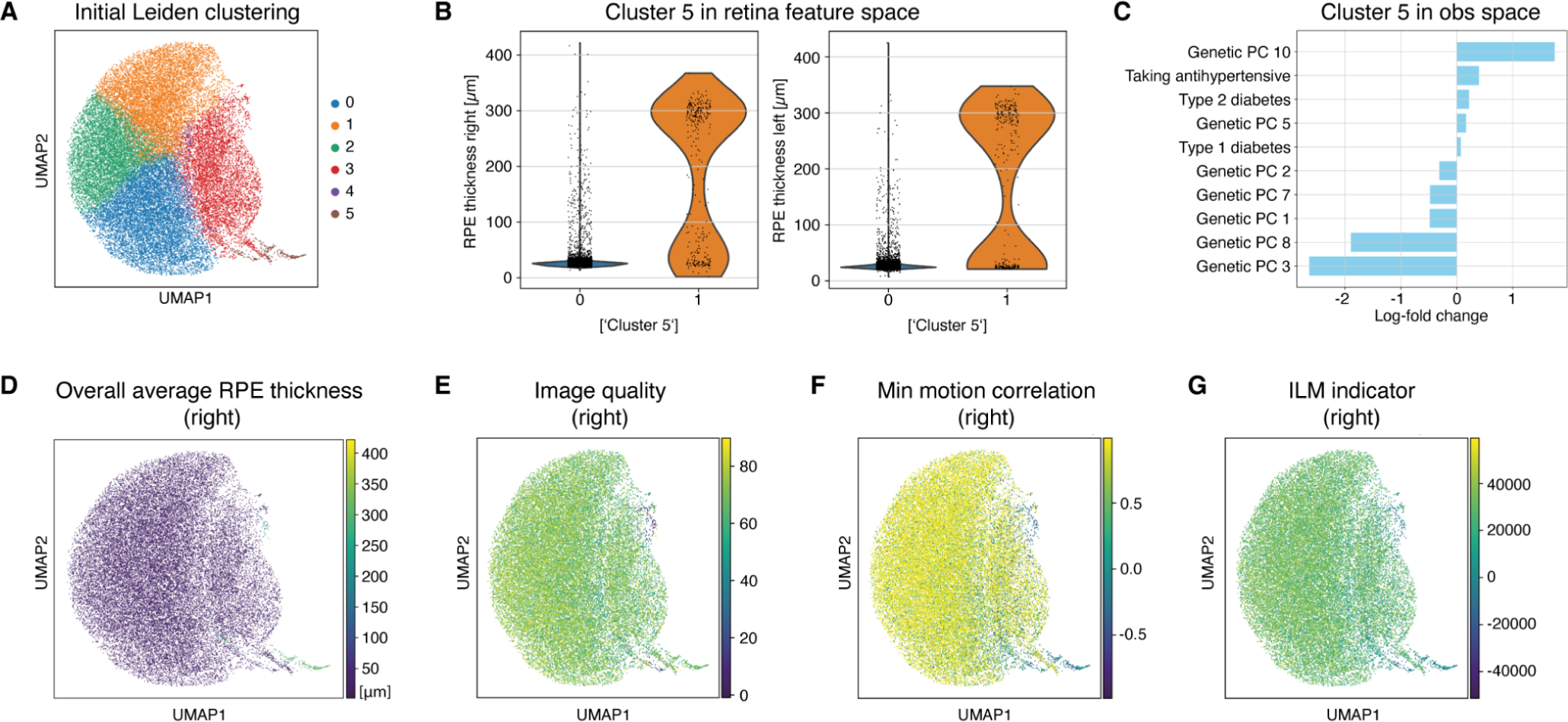
UK-Biobank retina derived feature quality control. (A) Leiden Clustering of retina derived feature space. (B) Comparison of ‘overall retinal pigment epithelium (RPE) thickness’ values between cluster 5 and the rest of the population. (C) Log ratio of top and bottom 5 fields in obs dataframe between cluster 5 and the rest of the population. (D) RPE thickness in the right eye outliers on the UMAP largely corresponds to cluster 5. (E) Image Quality of the OCT scan as reported in the UKB. (F) Minimum motion correlation correlation quality control indicator. (G) Inner limiting membrane (ILM) quality control indicator. (D-G) Data shown for the right eye only, comparable results for the left eye omitted.

**Supplementary Figure 8.**
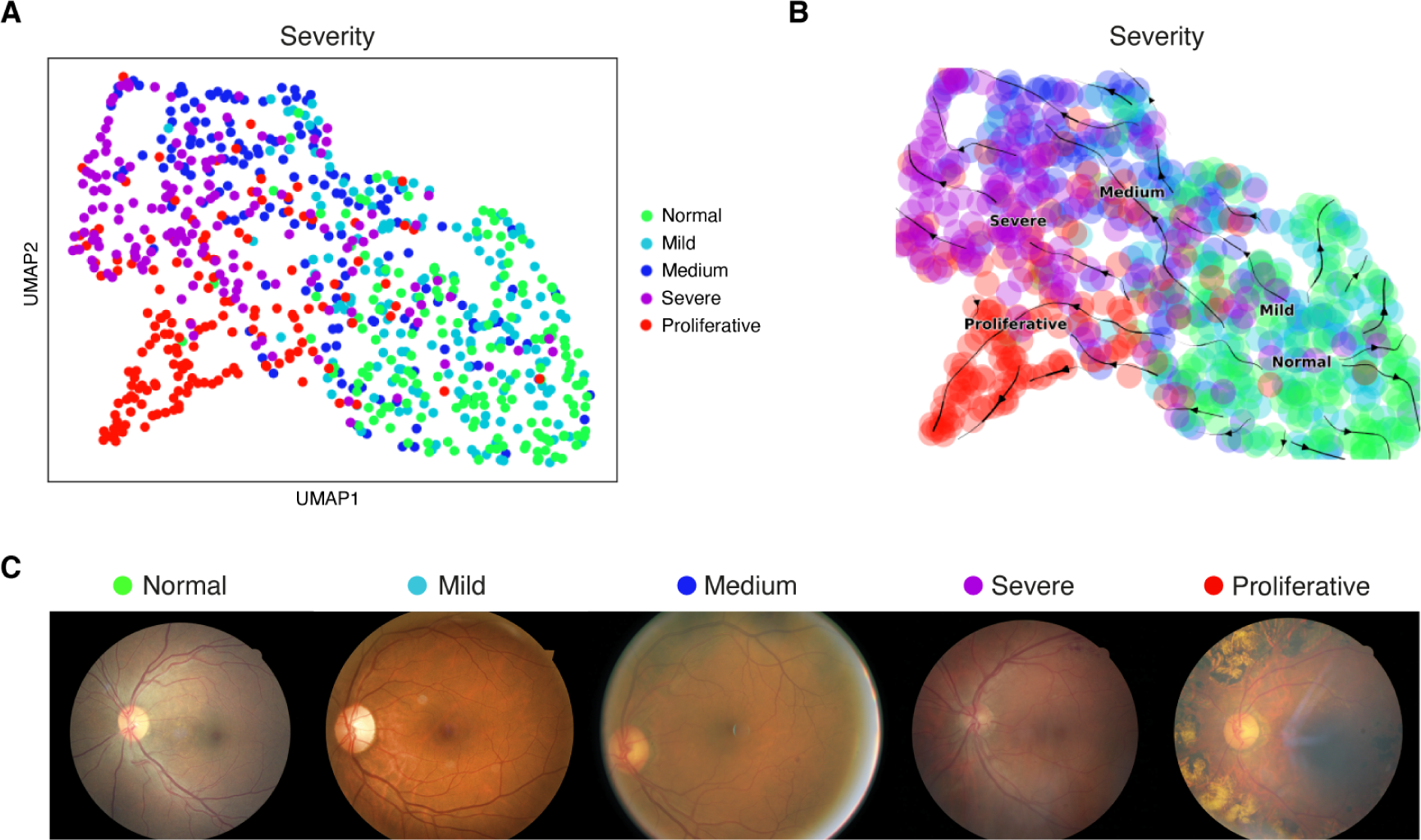
Recovery of diabetic retinopathy disease severity trajectory in human retina fundus photographs. Diabetic retinopathy, a complication of diabetes, specifically affects the eyes by causing damage to the blood vessels in the retina, the light-sensitive tissue at the back of the eye. Medical professionals utilize eye retina images, often obtained through techniques like fundoscopy, to diagnose and monitor diabetic retinopathy, assessing the extent of damage to the retina’s blood vessels and guiding treatment decisions. Using a pre-trained residual neural network, we generated a latent embedding (A) using ehrapy that we subsequently used for fate mapping to unveil disease severity trajectories in diabetic retinopathy (B). Patients previously labeled as “Normal” are expected to stay within that group. Our workflow was further able to clearly separate “Severe” and “Proliferative” cases into distinct clusters with unique trajectories. (C) Example retina fundus photography images of the retinopathy dataset.

